# Interictal functional ultrasound imaging in human epilepsy resection and responsive neurostimulation surgery

**DOI:** 10.64898/2026.10.15.26365563

**Authors:** Kofi A. Agyeman, Darrin J. Lee, Brian Lee, Ryan J. McGinn, Aidin Abedi, Jonathan Russin, Kevin Wu, Wooseong Choi, Charles Liu, Vassilios N. Christopoulos

## Abstract

Over 30% of patients with epilepsy are refractory to medication and may require surgical or neuromodulation-based intervention. Accurate localization of seizure-onset zones (SOZ) and assessment of treatment-associated brain responses remain major clinical challenges. Current workflows may involve invasive intracranial monitoring, prolonged hospitalization, adjustment of anti-seizure medications, and seizure capture to localize the SOZ, while opportunities for complementary intraoperative functional assessment remain limited. We used intraoperative functional ultrasound imaging (fUSI) to characterize interictal (i.e., non-seizure) cortical activity in nine patients with drug-resistant focal epilepsy and to assess treatment-associated brain responses following SOZ resection and responsive neurostimulation (RNS). fUSI signals were acquired directly from the exposed cortex during standard-of-care surgery. We developed six ultrafast power Doppler-based interictal activity markers (IaMs) to distinguish SOZ from peri-resection zone (PRZ) cortex and to quantify treatment-associated changes. The IaMs revealed distinct interictal hemodynamic dynamics between SOZ and PRZ cortex. Following SOZ resection, all six markers changed significantly within the surrounding PRZ. RNS also induced robust, stimulation paradigm-dependent modulation of cortical hemodynamic dynamics. Notably, RNS and surgical resection produced concordant directional changes across five of the six IaMs, despite representing fundamentally different therapeutic interventions. Together, these findings demonstrate the feasibility of intraoperative fUSI for characterizing and distinguishing interictal dynamics within SOZ and PRZ cortex and assessing treatment-associated cortical responses during human epilepsy surgery. Furthermore, fUSI could provide a complementary high-resolution functional imaging approach alongside established clinical methods for SOZ localization and intraoperative functional assessment.

## Introduction

Over seventy million people worldwide live with epilepsy, imposing substantial health, psychological, and economic burdens ^1–3^. Recent systematic reviews estimate the annual economic impact of epilepsy to exceed $50 billion in the United States and $120 billion globally ^4–6^. Approximately 30% of patients remain refractory to antiepileptic drug treatment ^7–9^ and may require surgical ^10,11^ or neuromodulation-based intervention^12^. Although surgical resection remains the gold standard for long-term seizure control in drug-resistant epilepsy^10,12,13^, its effectiveness critically depends on accurate localization of the seizure-onset zone (SOZ). Despite advances in neuroimaging and presurgical evaluation, reliable localization and prediction of postoperative outcomes remain challenging ^14,15^. Current clinical workflows integrate multimodal non-invasive evaluation with invasive intracranial monitoring when needed to localize SOZs and determine its relationship to functionally critical brain regions. Stereoelectroencephalography (sEEG) provides three-dimensional electrophysiological sampling of deep and superficial brain networks, although spatial sampling is constrained by electrode coverage. Intraoperative electrocorticography (iopECoG) provides complementary electrophysiological information during surgery, although its ability to predict postoperative seizure freedom remains variable^16–19^. Thus, there is a critical need for high-resolution, quantitative approaches that can characterize SOZ during the interictal state and assess the functional effects of resection intraoperatively, while complementing established clinical methods.

When resection is not feasible, particularly when seizures arise from eloquent cortex or patients are poor surgical candidates, neurostimulation therapies provide an alternative treatment strategy ^20,21^. Among these approaches, responsive neurostimulation (RNS) provides targeted electrical stimulation in response to detected epileptiform activity and is an established therapy for drug-resistant focal epilepsy ^20,22–25^. However, the acute effects of RNS on local brain activity remain incompletely characterized, in part because stimulation-evoked brain responses are difficult to assess directly ^20–25^. Intraoperative functional imaging could therefore provide a unique opportunity to characterize RNS-evoked cortical responses at the time of implantation.

Functional ultrasound imaging (fUSI) provides an opportunity to address these unmet intraoperative needs by enabling high-resolution imaging of cerebral hemodynamics during surgery. fUSI is an emerging neuroimaging technique that uses ultrafast ultrasound acquisition to measure changes in cerebral blood volume (CBV) as an indirect hemodynamic correlate of neuronal activity. Compared with other hemodynamic imaging techniques, fUSI offers a unique combination of high spatiotemporal resolution (∼100 μm spatially and up to 10 ms temporally), sensitivity to slow blood flow (down to ∼1 mm/s), and a large imaging depth (up to ∼6 cm) ^26,27^. Although adult human brain fUSI generally requires craniotomy ^28^ or an acoustic window because the skull attenuates and distorts ultrasound waves,^32,33^ it is particularly well suited to intraoperative functional assessment when surgical exposure already provides acoustic access to the brain. fUSI has successfully resolved large-scale neurovascular activity in both preclinical and clinical settings ^26,28–30^, making it a promising approach for evaluating epilepsy-related functional alterations and treatment-associated brain responses.

To date, applications of fUSI in epilepsy have largely focused on ictal or ictal-interictal states ^29–32^, providing important insights into SOZ localization and seizure propagation dynamics. However, whether interictal fUSI signals can provide quantitative information about clinically defined SOZ in humans remains unclear. Here, we use intraoperative fUSI to characterize interictal neuronal activity in anesthetized patients undergoing surgical resection or RNS implantation for drug-resistant focal epilepsy. Using direct acoustic access provided during standard-of-care epilepsy surgery, we acquire interictal fUSI signals and develop six ultrafast power Doppler-based interictal activity markers (IaMs) that distinguish SOZ from surrounding PRZ cortex. We further characterize changes in these markers within PRZs following SOZ resection and evaluate cortical responses to three clinically applied RNS stimulation paradigms. Together, these findings establish intraoperative fUSI as a complementary, high-resolution approach for characterizing interictal hemodynamic dynamics within the SOZ and assessing treatment-associated cortical responses during human epilepsy surgery.

## Results

To investigate cortical hemodynamic signatures associated with drug-resistant focal epilepsy and to examine the effects of surgical resection of SOZ, we recruited six (N=6) patients (3 females and 3 males; age range, 27 – 51 years) undergoing resection of presumptive epileptogenic regions under general anesthesia. Prior to surgery, patients underwent standard Phases I and II clinical evaluations, including interictal and ictal assessments, to localize the SOZ and define the planned resection margins (i.e., peri-resection zone). For fUSI analysis, SOZ (Fig. 1A, red dots) and surrounding PRZ were defined according to the clinical localization and planned surgical margins established through multimodal presurgical assessment. fUSI regions of interest corresponding to the SOZ and PRZ cortex were defined based on seizure-onset sites (red dots) and planned surgical margins (blue lines), as determined by invasive sEEG according to standard clinical practice (Fig. 1A, representative patients, P1, P5). A miniaturized 15.6-MHz fUSI linear array transducer was inserted through a cranial window and positioned on the cortical surface to acquire power Doppler (pD) signals from SOZ and PRZ cortex during the interictal state, both before and after resection (Fig.1 B). In addition, intraoperative electrocorticography (iopECoG) was performed over the same cortical regions, providing complementary electrophysiological evidence supporting the preoperatively defined SOZ localization and PRZ cortex (Fig. 1B, representative patient, P5, 1C). Each fUSI acquisition lasted either 120 or 180 seconds (Fig. 1C), producing pD images with 100 μm x 100 μm in-plane spatial resolution, 400 μm slice thickness, and a field of view (FOV) of 12.8 mm x 9.1 mm. Representative pD images revealed clearly resolved cortical vascular anatomy within the SOZ before resection (pre-SOZ) and the surrounding peri-resection cortex both before (pre-resection, pre-PRZ) and after (post-resection, pre-PRZ) SOZ resection (Fig. 1D). Because the anatomical location of the SOZ differed across patients, fUSI acquisition sites were selected according to each patient’s clinically defined SOZ. However, all recordings were obtained from frontal or temporal cortical regions (P1, left temporal; P2, left frontal lobe; P3-P5, right temporal lobe; and P6, left frontal lobe/ frontal cortical dysplasia). Detailed patient clinical characteristics and assessments are provided in Table S1.

**Figure 1.**
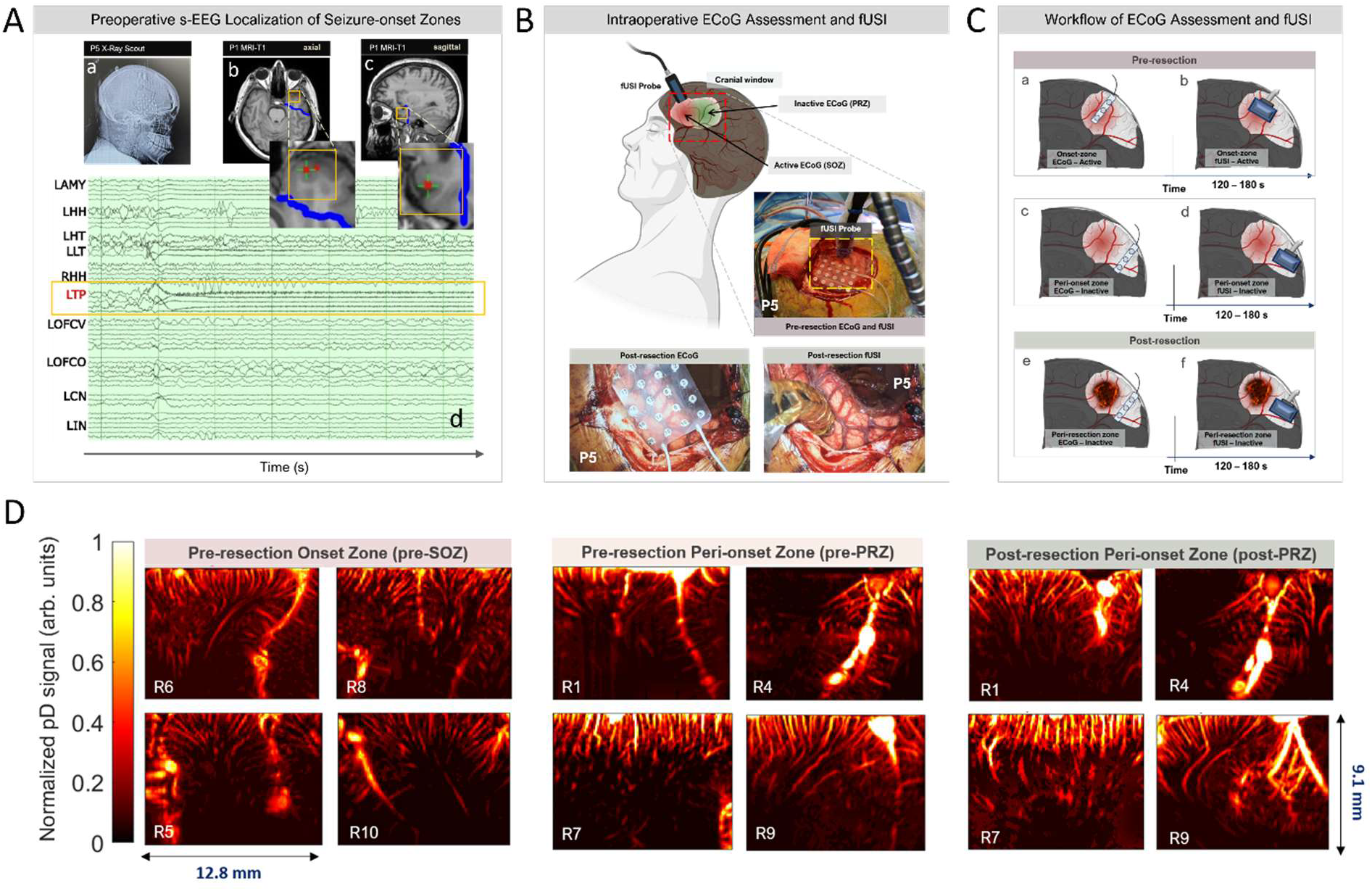
Experimental protocol for intraoperative fUSI mapping of interictal hemodynamic activity in the SOZ and PRZ cortex. **A)** Preoperative localization of SOZ and PRZ cortex. **a)** X-ray scout image showing sEEG electrodes in P5. **b)** Axial T1 weighted MR images centered on the SOZ. **c)** Sagittal T1 weighted MR images centered on the SOZ in Patient 1. Green makers, location of the displayed MRI slides; red dots, SOZ; blue line, planned resection margin. **d)** Full sEEG montage, excluding artifactual leads, demonstrating seizure-onset at the first two contacts of the left temporal pole (LTP) (yellow boxes). **B)** Intraoperative fUSI acquisition during standard-of-care epilepsy surgery under general anesthesia. Interictal signals were acquired through a cranial window using a transducer mounted on an articulating surgical arm to maintain stable positioning throughout each acquisition. Intraoperative images show representative locations of fUSI recordings on the exposed cortical surface. **C)** Schematic overview of the experimental protocol for acquiring interictal fUSI signals from the SOZ and surrounding PRZ cortex before and after SOZ resection. Each acquisition lasted 120 or 180 seconds. SOZ localization and planned resection margins were established during preoperative clinical evaluation, including sEEG and iopECoG providing complementary electrophysiological assessment. **D)** Representative pD images from the SOZ before resection (left panels), and adjacent PRZ cortex before (middle panels) and after (right panels) SOZ resection, with four acquisitions shown per condition. Images demonstrate clearly resolved cortical vascular anatomy within the imaged frontal and temporal cortical regions. ** L = Left; R = Right; AMY = amygdala; HH = hippocampal head, HT = anterior hippocampal tail, LT = lateral temporal extent of lead extending to LHT, TP = temporal pole, OFC = orbitofrontal cortex, V = vertical approach, O = orthogonal approach, CN = anterior cingulate, IN = anterior insula.

Because fUSI is a hemodynamic imaging technique, it does not provide an absolute measure of neuronal activity but instead quantifies relative changes in blood flow with respect to a baseline or reference state. In conventional functional neuroimaging paradigms, this reference is often defined by a resting condition, a control task, or a temporal baseline. During interictal fUSI recordings, however, such a reference is unavailable because SOZ and the surrounding PRZ exhibit fluctuating activity without a clearly defined resting state. Consequently, distinguishing the SOZ from surrounding PRZ and assessing changes before and after resection required analytical approaches that characterize intrinsic functional organization and local network dynamics directly from the ongoing interictal activity.

To address this challenge, we developed six quantitative markers derived from the pD signals (Fig. 2). Each pD image sequence, represented as a three-dimensional *space* × *time* data matrix, was spatially partitioned into non-overlapping rectangular regions (*dRs*; Fig. 2A-B). Within each dR, the pD signals from all pixels were spatially averaged to generate a single representative pD time series *x_i_* (*t*) for that region. Because these regional pD signals exhibited slow temporal trends and nonstationary fluctuations, each time series was modeled using an autoregressive integrated moving average (ARIMA) framework, yielding a model-predicted component 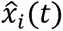 and a residual signal (i.e., 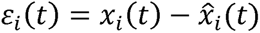), representing fluctuations not captured by the model. We then characterized both interactions among discretized regional signals and temporal properties of the regional signals. Local connectivity (LC) quantifies the local inter-regional synchronization by computing pairwise Pearson cross-correlation coefficients between the residual signals of the *dRs*, *ε_i_*(*t*), generating an LC matrix for each acquisition. The resulting LC matrices were then averaged across acquisitions to obtain the mean LC measure (Fig. 2A-b, -c). The remaining five markers characterized complementary aspects of the temporal dynamics of the regional pD signals and their model-derived representations.

**Figure 2.**
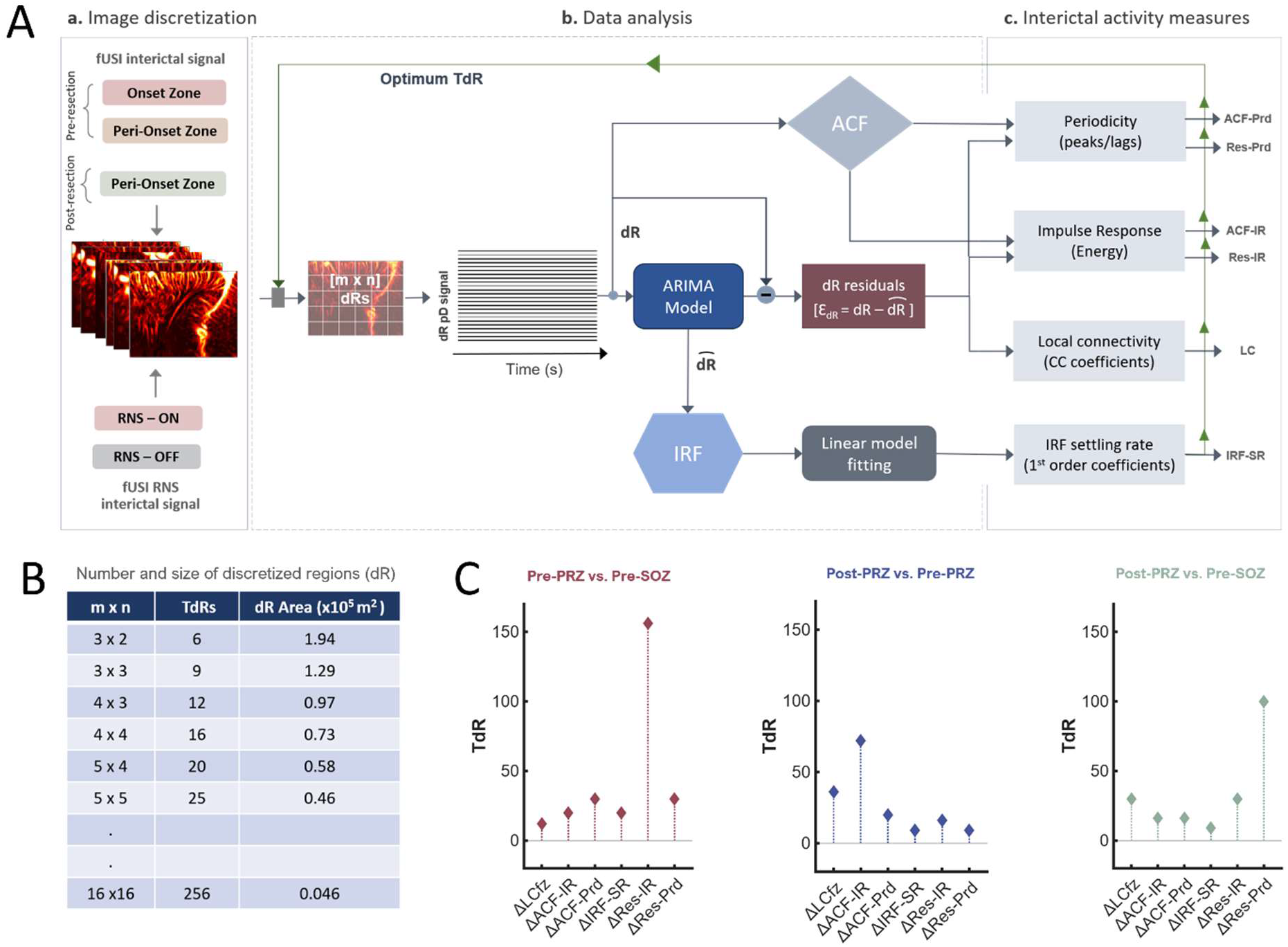
Analytical framework for deriving interictal activity markers (IaMs) from fUSI signals. **A)** Overview of the analysis workflow applied to interictal pD signals acquired from cortical seizure-onset zone SOZ, adjacent peri-resection regions (PRZ), and during responsive neurostimulation (RNS). Six interictal activity markers (IaMs) were derived: autocorrelation function impulse response (ACF-IR), autocorrelation function periodicity (ACF-Prd), pD residual impulse response (Res-IR), pD residual periodicity (Res-Prd), local connectivity (LC), and impulse response function settling rate (IRF-SR). Each fUSI imaging field of view (FOV) was partitioned into non-overlapping discretized regions (dRs), from which regional pD time series were extracted and used to derive the IaMs. **B)** Spatial discretization configurations evaluated for IaM computation. Twenty-eight dR configurations, ranging from *3* × *2* to *16* × *16* grids, were evaluated, corresponding to 6-256 total discretized regions (TdRs) and individual dR areas ranging from 1.94 to 0.046 mm^2^. **C)** Selection of the optimum spatial discretization for each IaM. For each experimental comparison, the optimum TdR was defined as the discretization yielding the largest mean difference in the corresponding marker (ΔIaM), averaged across recordings and dRs. Comparisons included pre-resection SOZ (pre-SOZ) versus pre-resection peri-resection zone (pre-PRZ; red stems), pre-PRZ versus post-resection PRZ (post-PRZ; blue stems), and pre-SOZ versus post-PRZ (green stems).

We first quantified the magnitude of fluctuations in each residual time series *ε_i_*(*t*), by calculating the area under the curve, yielding the pD residual impulse response (Res-IR) measure. Res-IR reflects the cumulative magnitude of residual fluctuations over time, providing an energy-like measure of ongoing local hemodynamic dynamics after removal of slow temporal trends. To assess the stability of the ARIMA model 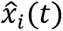, we applied a unit impulse input (i.e. Dirac delta function) to the corresponding transfer function of the time-series and evaluated the settling behavior of the resulting impulse response function (IRF) over time, up to 1000 s after perturbation. We fitted a linear model to approximately the final two-thirds (66.7%) of the IRF curve and defined the first-order coefficient (slope) as the impulse response function settling rate (IRF-SR) (Fig. 2A-b, -c). We hypothesized that IRF-SR reflects the ability of the ARIMA model 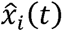 to capture temporal dynamics of the regional hemodynamic signal, which may depend on the degree of local synchrony within a *dR* (see details in the Methods section). We additionally computed the autocorrelation function (ACF) for each regional time series *x_i_* (*t*) across 116 lag steps to characterize temporal linear dependencies and oscillatory features of the interictal dynamics. From each ACF, we derived two measures: (i) ACF-IR, defined as the area under a fitted ACF curve and reflecting the strength of temporal dependence; and (ii) ACF-Prd, a periodicity index defined by the frequency (cycles per 1 s lag step) at which the ACF coefficients alternate between local maxima and minima, capturing oscillatory structure (Fig. 2A-b, -c).

### Interictal dynamics differentiate seizure-onset zone (SOZ) and peri-resection zone (PRZ) before resection

We acquired fUSI signals from the SOZ and adjacent PRZ to investigate differences in the intrinsic hemodynamic profiles between presumptive epileptogenic tissue and surrounding unresected cortex (Fig. 1B, C). Prior to SOZ resection, we acquired a total of twenty-one (n=21) pD recordings from SOZ (pre-SOZ; N=5, n=11) and peri-resection zone (pre-PRZ, N=6, n=10), where *N* denotes the number of patients and *n* denotes the number of recordings (Fig. 1B, C). SOZ localization and planned resection margins were defined based on the integrated standard Phase I and Phase II clinical evaluations (Fig. 1A–C). These clinically defined SOZ and PRZ labels served as the ground truth for the subsequent fUSI analyses and for evaluating whether fUSI-derived markers could distinguish the SOZ from adjacent PRZ cortex.

To evaluate the six interictal activity markers for each SOZ and PRZ recording, we followed the analytical framework outlined in Fig. 2. Each fUSI recording was partitioned into an *m* x*n* grid of non-overlapping dRs, and the pD signals from all pixels within each dR were spatially averaged to generate a representative regional pD time series. To determine the spatial discretization most sensitive to differences in each IaM between experimental conditions, we systematically evaluated 28 grid configurations, ranging from *3* × *2* to *16* × *16* grids (corresponding to 6-256 total dRs), with individual dR areas ranging from 1.94-0.046 mm^2^ per region (Fig. 2B). This systematic analysis allowed us to examine how each IaM varied as a function of spatial discretization (i.e., dR size) for three different comparisons: pre-resection SOZ (pre-SOZ) versus pre-resection PRZ (pre-PRZ), pre-PRZ versus post-resection PRZ (post-PRZ) and pre-SOZ versus post-PRZ (Fig. S1A-F). This approach allowed us to identify, for each IaM and experimental comparison, the spatial discretization that maximized the mean difference in the corresponding marker (ΔIaM), thereby defining the optimum dR area and total number of dRs (Fig. 2C).

The results showed that for the pre-resection SOZ versus PRZ comparison, the optimum spatial discretization (i.e., the configuration yielding the largest mean ΔIaM among dR sizes evaluated) correspond to 12, 20, 30, 20, 156, and 30 for the LC, ACF-IR, ACF-Prd, IRF-SR, Res-IR and Res-Prd markers, respectively (Fig. 2C, red stems). Using *Student’s t-test* analysis at these optimum discretization levels, we found significantly lower values for several IaMs in pre-PRZ compared with the pre-SOZ regions. For LC, Fig. 3A shows the mean Pearson cross-correlation coefficient matrices computed from the pD residual signals at the optimum TdR and averaged across recordings for the pre-SOZ and pre-PRZ conditions. The mean LC was significantly higher in pre-SOZ than in the pre-PRZ, with mean Fisher *z-*transformed correlation coefficients of 1.06 ± 0.023 (mean ± s.e.m; bottom-left triangle) and 0.79 ± 0.030 (top-right triangle), respectively (p < 1.0x10^-6^, Fig. 3B).

**Figure 3.**
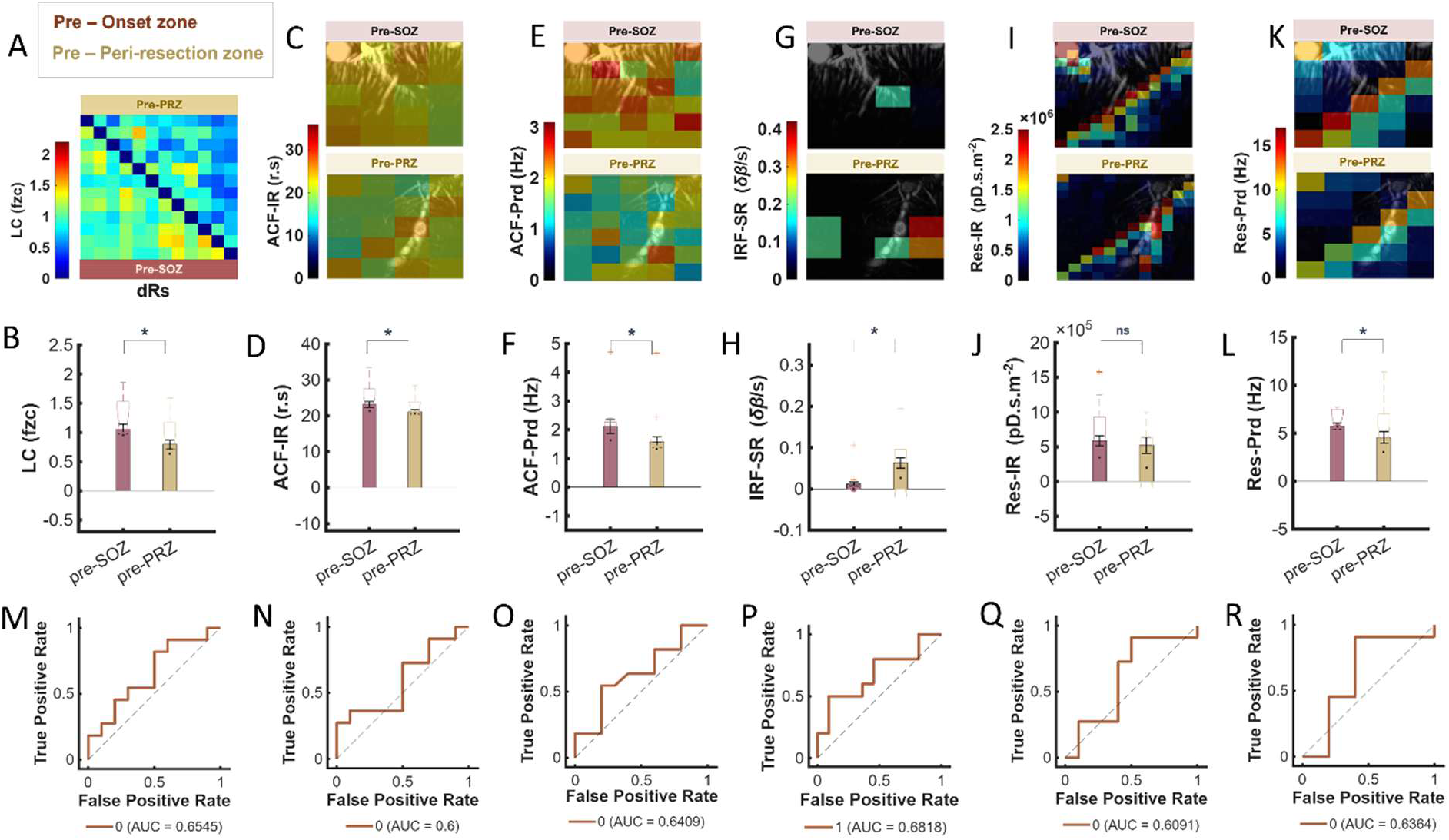
Interictal activity markers (IaMs) differentiate SOZ and PRZ zones prior to resection. **A)** Mean Pearson cross-correlation coefficient matrix computed from pD residuals at the optimum discretization for the LC measure (TdR = 12) and averaged across recordings. Correlations within the seizure-onset zone (pre-SOZ) are shown in the lower-left triangle (red) and those within the peri-resection zone (pre-PRZ) in the upper-right triangle (yellow). **B)** Mean local connectivity (LC) derived from Pearson cross-correlation coefficients, showing higher values in pre-SOZ than pre-PRZ (p < 1.0x10^-6^). **C)** Mean autocorrelation function impulse response (ACF-IR) for each dR in preSOZ (top) and pre-PRZ (bottom), averaged across recordings at the optimum discretization (TdR = 20) and overlaid on representative grayscale, mean pD vascular maps. ACF-IR represents the area under the ACF waveforms obtained by fitting the autocorrelation function computed over 116 lag steps. **D)** Mean ACF-IR across dRs and recordings, showing a significant difference between pre-SOZ and pre-PRZ (p < 1.0x10^-2^). **E)** Mean autocorrelation function periodicity (ACF-Prd) for each dR in pre-SOZ (top) and PRZ (bottom), averaged across recordings at the optimum discretization (TdR = 30) and overlaid on representative grayscale, mean pD vascular maps. ACF-Prd represents cycles per lag step derived from peak-to-trough oscillations in dR ACF waveforms over 116 lag steps. **F)** Mean ACF-Prd across dRs and recordings, showing significant difference between pre-SOZ and pre-PRZ (p < 1.0x10^-3^). **G)** Mean impulse response function settling rate (IRF-SR) for each dR in pre-SOZ (top) and pre-PRZ (bottom), averaged across recordings at the optimum discretization (TdR = 20) and overlaid on representative grayscale, mean pD vascular maps. IRF-SR represents the slope derived from dR IRF waveforms evaluated up to 1000 s after perturbation. **H**) Mean IRF-SR across dRs and recordings, showing lower values in pre-SOZ than pre-PRZ (p < 0.05). **I)** Mean pD residual impulse response (Res-IR) for each dR in pre-SOZ (top) and pre-PRZ (bottom), averaged across recordings at the optimum discretization (TdR = 156) and overlaid on representative grayscale, mean pD vascular maps. **J)** Mean Res-IR across dRs and recordings, showing no significant difference between pre-SOZ and pre-PRZ (p = 0.55). **K)** Mean pD residual periodicity (Res-Prd) for each dR in pre-SOZ (top) and pre-PRZ (bottom), averaged across recordings at the optimum discretization (TdR = 30) and overlaid on representative grayscale, mean pD vascular maps. **L)** Mean Res-Prd across dRs and recordings, showing lower values in pre-PRZ than pre-SOZ (p *<* 0.05). **M)** Receiver operating characteristics (ROC) curve and corresponding area under curve value (AUC) for distinguishing pre-SOZ and pre-PRZ recordings using LC at the optimum TdR. **N)** Same as M for ACF-IR measure. **O)** Same as M for ACF-Prd measure. **P)** Same as M for IRF-SR measure. **Q)** Same as M for Res-IR measure. **R)** Same as M for Res-Prd measure.

For the ACF-based markers, spatial maps of ACF-IR and ACF-Prd values across dRs at their respective optimum TdRs, averaged across recordings and overlaid on representative cortical pD vascular maps, revealed lower values in the pre-PRZ (bottom row) than in the pre-SOZ (top row) (Fig. 3C, E). Mean ACF-IR was significantly lower in pre-PRZ (21.06 ± 0.52 pD·s) than in pre-SOZ (23.14 ± 0.49 pD·s; mean ± s.e.m., p < 1.0x10^-2^), indicating higher ACF-IR levels in tissue from the SOZ (Fig. 3D). Similarly, mean ACF-Prd was significantly lower in pre-PRZ (1.57 ± 0.09 Hz) than in pre-SOZ (2.12 ± 0.10 Hz; mean ± s.e.m., p < 1.0x10^-3^), indicating higher periodicity levels in SOZ (Fig. 3F). A similar difference was observed for pD residual periodicity (Res-Prd). Spatial maps of mean Res-Prd values across dRs at the optimum discretization, averaged across recordings and overlaid on representative pD vascular maps, showed lower residual periodicity in the pre-PRZ (bottom panel) than in the pre-SOZ (top panel) (Fig. 3K). Mean Res-Prd was significantly lower in the pre-PRZ (4.57 ± 0.40 Hz; mean ± s.e.m.) than in pre-SOZ (5.75 ± 0.38 Hz; p < 0.05), indicating higher residual periodicity in SOZ (Fig. 3L). Together, these findings demonstrate that the LC, ACF-IR, ACF-Prd and Res-Prd markers consistently exhibited lower values in PRZs than in SOZs before resection.

In contrast, the mean IRF-SR maps shown in Fig. 3G revealed a different pattern between SOZ and PRZ. At the optimum discretization, IRF-SR values averaged across recordings for each dR were lower in the pre-SOZ (top panel) than in the adjacent pre-PRZ (bottom panel). Consistent with this spatial pattern, mean IRF-SR was significantly higher in the pre-PRZ (6.29x10^-2^ ± 2.37x10^-2^ δB/s; mean ± s.e.m) than in the pre-SOZ (1.20x10^-2^ ± 9.20x10^-3^, δB/s; p < 0.05; Fig. 3H).

The Res-IR maps, which reflect the accumulation of energy over time in the residual pD signals within each dR, are shown in Fig. 3I. At the optimum TdR, Res-IR values averaged across recordings showed no apparent difference between the pre-SOZ (top panel) and pre-PRZ (bottom panel). Consistent with this observation, the mean Res-IR did not differ significantly between pre-SOZ and pre-PRZ (p = 0.55; Fig. 3J), indicating comparable levels of residual signal energy accumulation between the two regions (i.e, SOZ and PRZ) during the interictal state.

To examine the specificity and sensitivity of each activity marker in distinguishing local interictal activity of SOZ from surrounding PRZ, we generated receiver operating characteristics (ROC) curves for each IaM at its corresponding optimum TdRs (Fig. 3M – R). The areas under the ROC curves (AUC) were 0.65, 0.6, 0.64, 0.68, 0.61 and 0.64 for the LC, ACF-IR, ACF-Prd, IRF-SR, Res-IR and Res-Prd markers, respectively, indicating the capability of the IaMs to differentiate between fUSI interictal recordings from SOZ and adjacent PRZ. Among the evaluated markers, IRF-SR exhibited the highest discriminative performance (AUC = 0.68). Note that the moderate AUC values may reflect the spatially distributed nature of epileptogenic network activity and the functional coupling between SOZ and adjacent PRZ, commonly observed in epileptogenic brains.

Finally, to assess the robustness of these findings across spatial scales, we examined how variations in dR size influenced the direction and consistency of all IaMs. Although the optimum dR size varied across markers and comparisons, the overall trends were largely preserved across the range of dR sizes tested (Fig. S1A-F). Specifically, pre-resection signals from PRZ exhibited consistently lower LC, ACF-IR, ACF-Prd, Res-IR and Res-Prd values than those from SOZ, whereas IRF-SR was generally higher in the PRZ (Fig. S1A-F, red curves). Shaded regions represent the standard error of mean across TdRs and recordings. An exception was observed for IRF-SR at the smallest discretized region sizes (TdR < 225), where the direction of the difference reversed, with lower IRF-SR values in PRZs than in SOZs (Fig. S1D, red curve). This reversal may reflect the reduced signal-to-noise ratio associated with very small dRs, where averaging fewer pixels increases the influence of noise in the pD signal. Overall, these analyses indicate that the observed differences between PRZ and SOZ are robust to the choice of spatial discretization, except at the smallest spatial scales where reduced signal averaging can affect specific measures.

### Seizure-onset zone (SOZ) resection modulates interictal activity in peri-resection zone (PRZ)

The ability to characterize and quantify how resection of the SOZ alters cortical functional dynamics has substantial clinical relevance, particularly for guiding intraoperative decisions and predicting postoperative outcomes in epilepsy surgery. To examine these effects, we analyzed fUSI-derived signals acquired from cortical areas in PRZ before and after resection (Fig. 1B, C). In total, 20 recordings were obtained from six patients (N=6) with drug-resistant focal epilepsy who underwent intraoperative resection under general anesthesia. For each PRZ, recordings were acquired before and after SOZ resection (n = 10 per condition).

Following the same preprocessing steps described above, each fUSI recording was discretized into non-overlapping rectangular dRs, which were then used to compute the six IaMs (Fig. 2A). As above, 28 distinct dR configurations were evaluated (Fig. 2B). When comparing interictal activity in PRZ before and after SOZ resection, we found consistent trends across dR sizes. Specifically, LC, Res-IR, Res-Prd, and ACF-Prd decreased following resection, whereas IRF-SR and ACF-IR increased (Fig. S1A-F, blue curves). Importantly, the direction of change for each marker remained consistent across the full range of dR sizes tested.

Evaluation across the tested TdR configurations identified the largest mean differences between pre-PRZ and post-PRZ conditions in the IaMs at TdR values 36, 72, 20, 9, 16, and 9 for LC, ACF-IR, ACF-Prd, IRF-SR, Res-IR and Res-Prd respectively (Fig. 2C, blue stems). Using *Student’s t-test* at these optimum TdR configurations, we identified statistically significant post-resection changes in all six markers within the PRZ. The mean LC matrix, obtained by averaging LC matrices across recordings at the optimum TdR, revealed reduced Pearson cross-correlation coefficients among pD residuals following resection (Fig. 4A). Higher coefficients were observed pre-resection (pre-PRZ; lower-left triangle, yellow) than after resection (post-PRZ; upper-right triangle, green). Quantitatively, mean LC was significantly lower post-resection (post-PRZ: 0.39 ± 0.007; mean ± s.e.m., Fisher z-score) than pre-resection (pre-PRZ: 0.75 ± 0.009), indicating reduced local synchrony of neurovascular dynamics following SOZ resection (p < 1.0x10^-6^, Fig. 4B).

**Figure 4.**
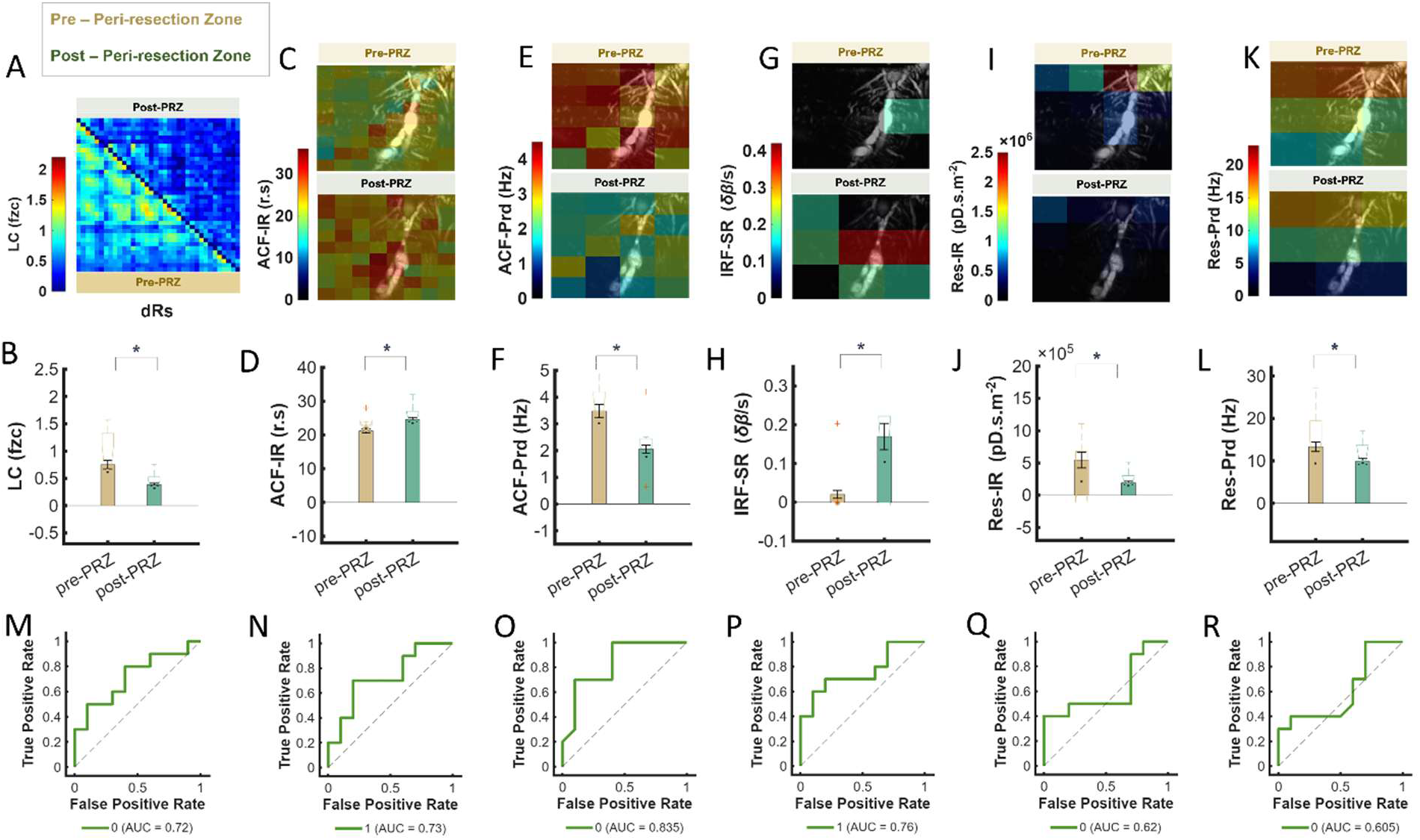
Interictal activity markers (IaMs) in PRZ before and after SOZ resection. **A)** Mean Pearson cross-correlation matrix computed from pD residuals at the optimum discretization for the local connectivity (LC) measure (TdR=36) and averaged across recordings. Correlations within the pre-PRZ are shown in the lower-left triangle (yellow) and those within the post-PRZ in the upper-right triangle (green). **B)** Mean LC, derived from Pearson cross-correlation coefficients, showing lower values in post-PRZ than pre-PRZ, yellow (p < 1.0x10^-6^). **C)** Mean autocorrelation function impulse response (ACF-IR) for each dR in pre-PRZ (top) and post-PRZ (bottom), averaged across recordings at the optimum discretization (TdR = 72) and overlaid on representative grayscale, mean pD vascular maps. ACF-IR represents the area under ACF waveforms obtained by fitting the autocorrelation functions computed over 116 lag steps. **D)** Mean ACF-IR across dRs and recordings, showing significant higher values in post-PRZ than pre-PRZ (p < 1.0x10^-6^). **E)** Mean autocorrelation function periodicity (ACF-Prd) for each dR in pre-PRZ (top) and post-PRZ (bottom), averaged across recordings at the optimum discretization (TdR = 20) and overlaid on representative grayscale, mean pD vascular maps. **F)** Mean ACF-Prd across dRs and recordings, showing significantly lower values in post-PRZ, than pre-PRZ (p < 1.0x10^-6^). **G)** Mean impulse response function settling rate (IRF-SR) for each dR in pre-PRZ (top) and post-PRZ (bottom), averaged across recordings at the optimum discretization (TdR = 9) and overlaid on representative grayscale, mean pD vascular maps. **H**) Mean IRF-SR across dRs and recordings, showing significantly higher values in post-PRZ than pre-PRZ (p < 0.05). **I)** Mean pD residual impulse response (Res-IR) for each dR in pre-PRZ (top) and post-PRZ (bottom), averaged across recordings at the optimum discretization (TdR = 16) and overlaid on representative grayscale, mean pD vascular maps. **J)** Mean Res-IR across dRs and recordings, showing significantly lower values in post-PRZ than pre-PRZ (p < 1.0x10^-2^). **K)** Mean pD residual periodicity (Res-Prd) for each dR in pre-PRZ (top) and post-PRZ (bottom), averaged across recordings at the optimum discretization (TdR = 9) and overlaid on representative grayscale, mean pD vascular maps. **L)** Mean Res-Prd across dRs and recordings, showing significantly lower values in post-PRZ than pre-PRZ (p < 1.0x10^-2^). **M)** Receiver operating characteristics (ROC) curve and corresponding area under curve (AUC) for distinguishing pre-PRZ and post-PRZ recordings using LC at the optimum TdR. **N)** Same as M for ACF-IR measure. **O)** Same as M for ACF-Prd measure. **P)** Same as M for IRF-SR measure. **Q)** Same as M for Res-IR measure. **R)** Same as M for Res-Prd measure.

ACF-Prd showed a similar reduction following resection. Mean ACF-Prd for each dR, averaged across recordings at the optimum TdR and overlaid on representative mean pD vascular maps, was lower in post-PRZ than pre-PRZ regions (Fig. 4E). Mean ACF-Prd across dRs and recordings was significantly lower post-resection (2.06 ± 0.13 Hz; mean ± s.e.m.) than pre-resection (3.49 ± 0.15 Hz.; p < 1.0x10^-6^), indicating reduced periodicity of local interictal pD dynamics following SOZ resection (Fig. 4F). Res-IR was also reduced following resection, with lower values for individual dRs in post-PRZ than pre-PRZ regions (Fig. 4I). Mean Res-IR across dRs and recordings was significantly lower post-resection (1.66×10□ ± 3.05×10□ pD·s·m⁻²; mean ± s.e.m.) than pre-resection (5.12×10□ ± 1.65×10□ pD·s·m⁻²; p < 1.0x10^-2^), indicating a significant reduction in residual impulse-response energy following SOZ resection (Fig. 4J). Similarly, Res-Prd values for each dR were lower in post-PRZ than pre-PRZ regions (Fig. 4K), with mean Res-Prd across dRs and recordings significantly lower post-resection (9.87 ± 0.83) than pre-resection (13.32 ± 1.00 Hz; mean ± s.e.m.; p < 1.0x10^-2^) (Fig. 4L). Together, the reductions in LC, ACF-Prd, and Res-Prd indicate decreased local synchrony and periodicity of interictal neurovascular dynamics following SOZ resection. The concurrent reduction in Res-IR further indicates a decrease in the magnitude and persistence of residual pD dynamics in the surrounding PRZ.

Conversely, IRF-SR and ACF-IR increased following resection. Mean ACF-IR for each dR, averaged across recordings at the optimum TdR and overlaid on representative mean pD vascular maps, was higher in post-PRZ than pre-PRZ regions (Fig. 4C). Quantitatively, mean ACF-IR across dRs and recordings was significantly higher post-resection (24.57 ± 0.32 r.s; mean ± s.e.m.) than pre-resection (21.17 ± 0.27 r.s; p < 1.0x10^-6^), indicating increased autocorrelation impulse-response energy following SOZ resection (Fig. 4D). Likewise, IRF-SR values for individual dRs were higher (i.e., steeper rise) in post-PRZ than pre-PRZ (Fig. 4G). Mean IRF-SR across dRs and recordings was significantly higher post-resection (0.17 ± 0.057 δB/s; mean ± s.e.m.) than pre-resection (0.02 ± 0.019, δB/s; p < 0.05) (Fig. 4H). The lower IRF-SR observed prior to resection is consistent with our earlier hypothesis that ARIMA models capture a more constrained temporal regime, in which the influence of a transient input dissipates rapidly. Following resection, IRF-SR increases while local connectivity decreases, suggesting disruption of the previously synchronized dynamics. This shift indicates a transition toward a less constrained temporal regime. In other words, after removal of the SOZ, local connectivity decreases, suggesting a transition from an overly and pathologically synchronized state to a less constrained system which may be capable of normal function.

The IaMs also showed greater discriminatory performance for distinguishing pre-PRZ from post-PRZ recordings than observed for the pre-SOZ versus pre-PRZ comparison. ROC analysis performed at the corresponding optimum TdRs for each IaM yielded AUC values of 0.72, 0.73, 0.84, 0.76, 0.62 and 0.61 for the LC, ACF-IR, ACF-Prd, IRF-SR, Res-IR and Res-Prd, respectively (Fig. 4M-R). Among these markers, ACF-Prd showed the strongest discrimination between pre- and post-resection PRZ dynamics, with an AUC of 0.84. Together, these results demonstrate that SOZ resection produces measurable changes in the interictal dynamics of the surrounding PRZ that can be detected using multiple complementary IaMs.

To further assess the impact of resection on interictal activity, we compared IaMs derived from signals recorded from SOZ prior to resection (pre-SOZ) with those from PRZ after resection (post-PRZ). All six IaMs differed significantly between pre-SOZ and post-PRZ (p < 0.016, Fig. S2) with the direction of change remaining consistent across the range of dR sizes tested (Fig. S1, green curves). Notably, these changes closely mirrored those observed when comparing pre-PRZ and post-PRZ recordings. The optimum TdRs were 30, 16, 16, 9, 30, and 100 for LC, ACF-IR, ACF-Prd, IRF-SR, Res-IR, and Res-Prd IaMs, respectively, corresponding to the largest differences between pre-SOZ and post-PRZ conditions (Fig. 2C green stems). At these optimum TdRs, ROC analysis yielded AUC values of 0.85, 0.72, 0.76, 0.77, 0.81, and 0.76, for LC, ACF-IR, ACF-Prd, IRF-SR, Res-IR, and Res-Prd, respectively.

### Responsive neurostimulation (RNS) modulates interictal dynamics of cortical seizure-onset zones

Resective epilepsy surgery aims to achieve seizure control by removing brain regions capable of independently producing seizures. In contrast, RNS seeks to modulate pathological neural activity without removing seizure-generating tissue, although its mechanism of action remains poorly understood. Characterizing how RNS alters cortical dynamics and local neural network connectivity may therefore provide insight into its effects on epileptic networks. Here, we used intraoperative fUSI to examine changes in interictal hemodynamic dynamics and connectivity during RNS stimulation at the time of device implantation.

To examine how RNS modulates spatiotemporal hemodynamic dynamics and connectivity, we acquired four interictal pD recordings (n = 4) from three patients (N = 3) with drug-resistant focal epilepsy. As described above, all patients underwent standard-of-care Phase I and Phase II evaluations to localize SOZs and guide RNS lead placement (Fig. 5A). RNS electrode configurations were determined by clinical indications and included strip and/or depth electrodes. Patient 7 (P7) had strip electrodes placed anterior and posterior to focal cortical dysplasia (FCD), Patient 8 (P8) had hippocampal strip and depth electrodes, and Patient 9 (P9) had depth electrodes in the left anterior insula and interhemispheric strip electrodes. fUSI was performed over cortical regions within and adjacent to the SOZ and between the RNS leads, as defined by stereo-EEG recordings (Fig. 5A, representative patient, P7). During standard-of-care RNS implantation under general anesthesia, the ultrasound probe was positioned over the exposed cortex through the craniotomy (Fig. 5B).

**Figure 5.**
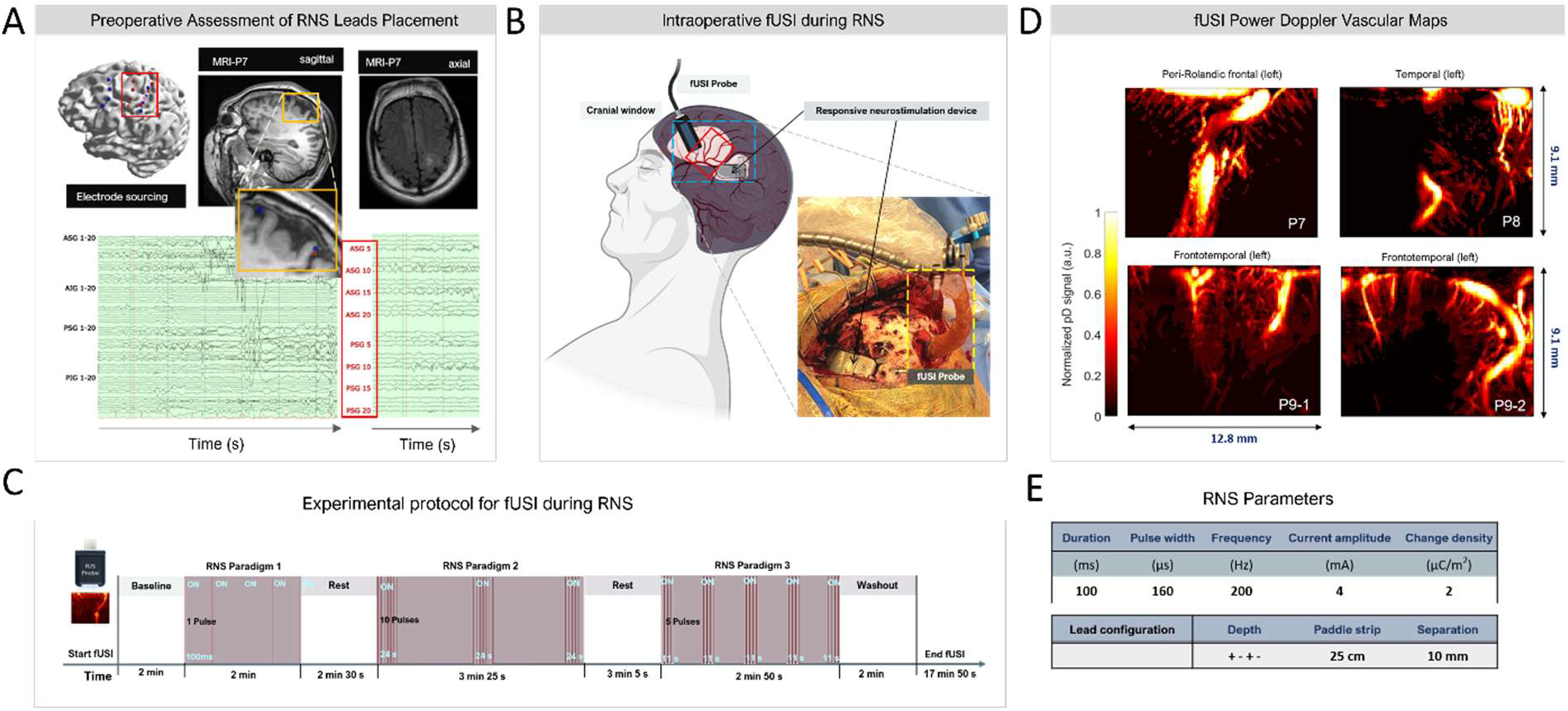
Responsive neurostimulation (RNS) electrode localization, intraoperative fUSI acquisition, and stimulation protocol. **A)** Preoperative localization of the SOZ and RNS leads in patient P7. Upper left: 3D model of patient’s brain showing the SOZ (red dots) and RNS strip electrodes (blue dots). Upper middle, sagital T1-weighted MRI. Upper right, axial T2-weighted MRI. MRI slices are centered on the lesional SOZ, indicated by the green marker. Lower panels, intracranial ECoG showing the full electrode montage consisting of four grids (left) and a limited montage (right). SOZ was broadly localized to ASG5/10/15/20 in the posterior portion of the anterior grid and PSG5/10/15/20 in the anterior portion of the posterior grid, corresponding to the locations indicated by red circles in the 3D reconstruction. **B)** Surgical workflow for standard-of-care implantation of an RNS device (NeuroPace RNS ® System, Model 320) in patients with drug-resistant focal epilepsy under general anesthesia. The fUSI transducer was mounted on a surgical articulating arm and positioned over the exposed cortical surface to acquire fUSI signals. C) Continuous fUSI acquisition during RNS stimulation, beginning with a 2-min baseline recording followed by three sequential RNS stimulation paradigms (Paradigms 1-3), with stimulation ON and OFF periods interleaved with rest periods, and concluding with a 2-min post-stimulation (i.e., washout) recording. **D)** Mean pD images showing cortical vascular anatomy within the fUSI FOV for patients P7, P8, and P9 during baseline recordings. Imaging fields included frontal and temporal cortical regions. **E)** RNS electrode configurations and stimulation parameters for each patient. All patients received identical stimulation settings consisting of pulses with a 160 µs pulse width, frequency of 200 Hz, charge density of 2 µC/m2 and 4 mA current amplitude. ** ASG = Anterior Superior Grid; PSG = Posterior Superior Grid; AIG: Anteroinferior grid; PIG = Posteroinferior grid.

fUSI was performed simultaneously with RNS using three sequential stimulation paradigms delivered during a 17-min and 50-s protocol. The protocol consisted of a 2-min baseline fUSI recording, followed by three RNS stimulation paradigms interleaved with rest periods and a final 2-min post-stimulation recording (Fig. 5C). All three patients received the same stimulation protocol using the FDA-approved NeuroPace RNS System (Model RNS-320 ®), with 160 µs pulse width, 200 Hz frequency, 2 µC/m^2^ charge density, and 4 mA current amplitude (Fig. 5E). Detailed parameters for each stimulation paradigm are provided in the Methods section. fUSI recordings included the left peri-Rolandic frontal cortex in P7, left temporal cortex in P8, and left frontotemporal cortex in P9 (Fig. 5D).

To characterize RNS-evoked effects on cerebral hemodynamics, we generated statistical parametric maps (SPMs) using a two-sided *Student’s t-test* (p<0.01) with false discovery rate (FDR) correction. The SPMs were computed through pixelwise comparisons of the mean pD signal recorded during RNS stimulation (pulses-ON) against reference signal acquired in the absence of stimulation (pulses-OFF). For each stimulation paradigm, SPMs were derived independently, using the mean pD signal from the 5 s immediately preceding each pulses-ON period as the reference activity (Fig 5C). For Paradigm 1, which consisted of a single 100 ms pulse, we analyzed 5 s of signal beginning at pulse onset. The resulting SPMs identified brain regions significantly modulated by each stimulation paradigm (Fig. 6A-C, P8, and Fig. S3, P7 and P9). These maps revealed robust, spatially localized changes in CBV within the FOV, with distinct cortical regions exhibiting CBV increases (red) or decreases (blue), overlaid on mean grayscale pD background images. In P8, clear CBV modulations were observed across all three paradigms (Fig. 6A-C, top panels). For visualization, only the top 5% of significantly modulated pixels, ranked by z-scores of CBV percentage change relative to the reference activity, are displayed.

**Figure 6.**
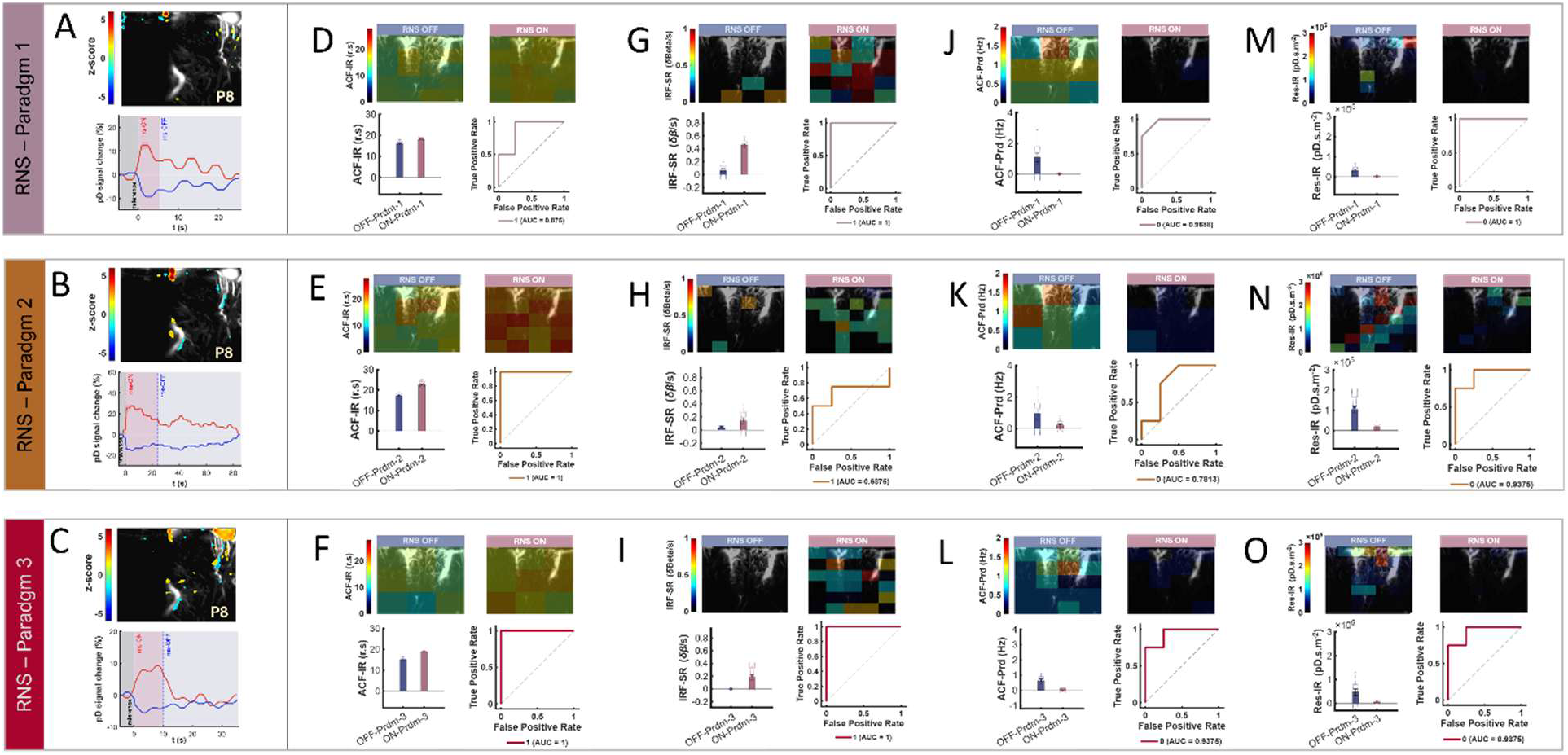
Statistical parametric maps (SPM), event-related average (ERA) waveforms of RNS-evoked hemodynamic responses, and stimulation-induced changes to interictal activity markers (IaMs). **A top panel)** SPM shows localized brain regions with significantly higher or lower CBV changes relative to the preceding 5 s of the stimulation pulses-OFF period used as reference activity (two-sided Student’s t-test, p < 0.01, FDR corrected for the number of pixels in the image) for RNS Paradigm 1 in P8. The SPM is overlaid on a grayscale, mean pD image from baseline. Only the top 5% of significantly modulated pixels, ranked by the z-scores of their percentage CBV changes relative to the reference activity, are displayed. **A bottom panel)** Corresponding ERA waveforms averaged across the top 5% of significantly modulated pixels identified in the SPM, showing CBV increases (red curve) or decreases (blue curve) during pulses-ON periods for RNS Paradigm 1. **B)** Same as panel A, for RNS Paradigm 2. **C)** Same as panel A, for RNS Paradigm 3. **D top panel)** Mean autocorrelation function impulse response (ACF-IR) for each dR during stimulation pulses-OFF (blue, left panel) and pulses-ON (red, right panel) periods, averaged across recordings at the optimum discretization (TdR = 20) for Paradigm 1 and overlaid on a representative mean pD grayscale vascular map. **D bottom left panel)** Mean ACF-IR (area under the ACF waveforms) across dRs and recordings showing significantly higher values during stimulation pulses-ON (red) than pulses-OFF (blue) periods (p < 1.0x10^-6^). **D bottom right panel)** Receiver operating characteristic (ROC) curve and corresponding area under the curve (AUC) generated for distinguishing RNS Paradigm 1 pulses-ON from pulses-OFF recordings using ACF-IR at the optimum TdR. **E)** Same as panel D for RNS Paradigm 2 (optimum TdR = 20, p < 1.0x10^-6^). **F)** Same as panels D for RNS Paradigm 3 (optimum TdR = 9, p < 1.0x10^-6^). **G top panel)** Mean impulse response function settling rate (IRF-SR) for each dR for RNS Paradigm 1 pulses-OFF (blue, left panel) and pulses-ON (right, right panel) periods, averaged across recordings at the optimum discretization (TdR = 20) and overlaid on a representative mean pD grayscale vascular map. **G bottom left panel)** Mean impulse response function settling rate (IRF-SR) across dRs and recordings, showing significantly higher values during stimulation pulses-ON (red) than pulses-OFF (blue) periods (p < 1.0x10^-6^). **G bottom right panel)** ROC curve and corresponding AUC for distinguishing RNS Paradigm 1 pulses-ON from pulses-OFF recordings using IRF-SR at the optimum TdR. **H)** Same as panel G for RNS Paradigm 2 (optimum TdR = 36, p < 1.0x10^-2^). **I)** Same as panel G for RNS Paradigm 3 (optimum TdR = 30, p < 1.0x10^-3^). **J top panel)** Mean autocorrelation function periodicity (ACF-Prd) for each dR during RNS Paradigm 1 stimulation pulses-OFF (blue, left panel) and pulses-ON (red, right panel) periods, averaged across recordings at the optimum discretization (TdR = 9) and overlaid on a representative mean pD grayscale vascular map. **J bottom left panel)** Mean ACF-Prd across dRs and recordings, showing significantly lower values during pulses-ON (red) than pulses-OFF (blue) periods (p < 1.0x10^-5^). **J bottom right panel)** ROC curve and corresponding AUC for distinguishing RNS Paradigm 1 pulses-ON from pulses-OFF recordings using ACF-Prd at the optimum TdR. **K)** Same as panel J for RNS Paradigm 2 (optimum TdR = 9, p < 1.0x10^-3^). **L)** Same as panel J for RNS Paradigm 3 (optimum TdR = 20, p < 1.0x10^-5^). **M top panel)** Mean pD residual impulse response (Res-IR) for each dR during RNS Paradigm 1 pulses-OFF (blue, left panel) and pulses-ON (red, right panel) periods, averaged across recordings at the optimum discretization (TdR = 36) and overlaid on a representative mean pD grayscale vascular map. **M bottom left panel)** Mean Res-IR across dRs and recordings, showing significantly lower values during pulses-ON (red) than pulses-OFF (blue) periods for Paradigm 1 (p < 1.0x10^-3^). **M bottom right panel)** ROC curve and corresponding AUC for distinguishing RNS Paradigm 1 pulses-ON from pulses-OFF recordings using Res-IR at the optimum TdR. **N)** Same as panel M for RNS Paradigm 2 (optimum TdR = 42, p < 1.0x10^-4^). **O)** Same as panel M for RNS Paradigm 3 (optimum TdR = 49, p < 0.05).

To evaluate the temporal dynamics of RNS-evoked activity, we generated event related average (ERA) waveforms. These waveforms represent CBV changes, expressed as percentage deviations from the mean reference activity during the 5 s preceding each pulses-ON period and averaged across the top 5% significantly modulated pixels (Fig. 6A-C, bottom panels, P8). The ERA curves capture the temporal profiles of hemodynamic responses associated with RNS stimulation. Across all three patients, the RNS paradigms elicited robust, regionally specific CBV changes. Notably, we identified brain regions where the ERA curves exhibited increasing (red curves) and decreasing (blue curves) trajectories during pulses-ON periods, with signals returning to baseline during pulses-OFF periods (Fig. 6A-C, bottom panels, P8). The corresponding SPM and ERA curves for the remaining patients are shown in Fig. S3, and a representative movie of cortical activation during RNS in P8 (Paradigm 3) is provided in Movie S1.

Next, we investigated the effects of RNS on interictal dynamics within SOZ by assessing stimulation-induced changes across the six IaMs developed in this study. We also compared the three RNS stimulation paradigms to assess paradigm-specific effects on the pD signal. pD signals acquired during pulses-ON and pulses-OFF periods were averaged across trials for each paradigm. To ensure balanced comparisons between the two conditions, pulses-ON signals were resampled using linear interpolation to match the number of samples in the corresponding pulses-OFF periods. The IaMs were then computed using the same processing pipeline described in Fig. 2. Specifically, the mean 3D matrices (*space* × *time*), derived by separately averaging the pD signals across pulses-ON and pulses-OFF periods, were discretized into *m* × *n* non-overlapping dRs. The resulting dR time-series signals were then used to compute the six IaMs for each RNS paradigm. Stimulation-induced changes in each IaM were quantified as the difference between its mean value during pulses-ON and pulses-OFF periods.

To assess the consistency and spatial-scale dependence of each IaM, we first evaluated how stimulation-induced changes varied as a function of dR size across all configurations. Specifically, we examined whether the direction and magnitude of the pulses-ON versus pulses-OFF differences were preserved across the full range of dR sizes for each RNS paradigm [Paradigm 1 (Prdm-1), Paradigm 2 (Prdm-2) and Paradigm 3 (Prdm-3)]. A total of 28 dR-size configurations were examined (Fig. 2B). This analysis revealed that four IaMs (ACF-IR, IRF-SR, Res-IR, and ACF-Prd) exhibited stable and consistent modulation during RNS stimulation across all dR-size configurations, indicating robustness to variations in spatial partitioning (Fig. S4B, C, D, E). In contrast, the LC and Res-Prd exhibited dR-dependent behavior, with the direction of stimulation-induced change varying across dR sizes and paradigms (Fig. S4A, F). Given this sensitivity to dR size, subsequent analyses focused on the four IaMs that demonstrated consistent, dR-independent modulation in response to RNS stimulation. We then identified the dR sizes that maximized the mean pulses-ON versus pulses-OFF differences for each of these four IaMs (Table S2A).

Using *Student’s t*-test analysis performed at these optimum dR sizes, we found that both ACF-IR (area under the ACF curve; Fig. 6D–F) and IRF-SR (slope of the IRF curve; Fig. 6G-I) were significantly increased during pulses-ON periods (ACF-IR: *p* < 1.00 × 10⁻^6^; IRF-SR: *p* < 0.004 across all three stimulation paradigms, bottom-left panels). The mean ACF-IR (Fig. 6D-F) and IRF-SR (Fig. 6G-I) values for each dR, averaged across recordings and overlaid on representative mean pD vascular maps, increased during RNS pulses-ON (top-right panels, red) compared with RNS pulses-OFF (top-left panels, blue) periods across all paradigms. In contrast, the mean ACF-Prd (Fig. 6J–L) and Res-IR (Fig. 6M–O) values for each dR, averaged across recordings decreased during RNS pulses-ON (top-right panels, red) compared with RNS pulses-OFF (top-left panels, blue) periods for all paradigms. Quantitatively, the ACF-Prd (cycle per 1 s lag step); Fig. 6J–L) and Res-IR (area under the residuals’ curves; Fig. 6M–O) were significantly decreased during pulses-ON periods (Res-IR: *p* < 0.018; ACF-Prd: *p* < 0.0006 across all three stimulation paradigms, bottom-left panels). Additionally, ROC analysis (Fig. 6D-O bottom-right panels) of ACF-IR, IRF-SR, ACF-Prd and Res-IR showed robust sensitivity and specificity in differentiating RNS-evoked effects on cortical interictal activity during pulses-ON and pulses-OFF periods, with the corresponding AUC values (Table S2B) summarizing the overall discriminatory performance of each marker.

Together, these findings indicate that RNS stimulation induces a systematic reorganization of local hemodynamic dynamics, characterized by increased short-range temporal integration and response sensitivity alongside reduced periodicity and residual autocorrelation during stimulation. The reductions in ACF-Prd and Res-IR may reflect decreased synchronous local activity or metabolic demand during stimulation. Furthermore, increased IRF-SR with RNS suggests stimulation evoked reduction in local activity synchrony, shifting the ARIMA dynamics toward a less tightly constrained temporal regime in which the influence of transient inputs persists over longer time scales.

Having identified IaMs that exhibit robust, dR-independent modulation and quantified their maximal effects at the optimum spatial scales, we next evaluated the overall impact of RNS stimulation by integrating responses across all spatial partition sizes, including IaMs that displayed dR-dependent behavior. This analysis allowed us to quantitatively assess differences among the three RNS paradigms while accounting for variation in dR-size. To this end, we assessed the net RNS-evoked effects by averaging the changes in each activity measure (ΔIaM) across all investigated TdRs (m=28; Fig. 2B). We then performed a two-way analysis of variance (ANOVA) with stimulation level (pulses-OFF and pulses-ON) and stimulation paradigm (Paradigm 1 (Prdm-1), Paradigm 2 (Prdm-2) and Paradigm 3 (Prdm-3)) as factors. The ANOVA revealed significant main effects of both stimulation condition and paradigm for all six IaMs (p < 0.03). Detailed statistics are provided in Fig. 7A.

**Figure 7.**
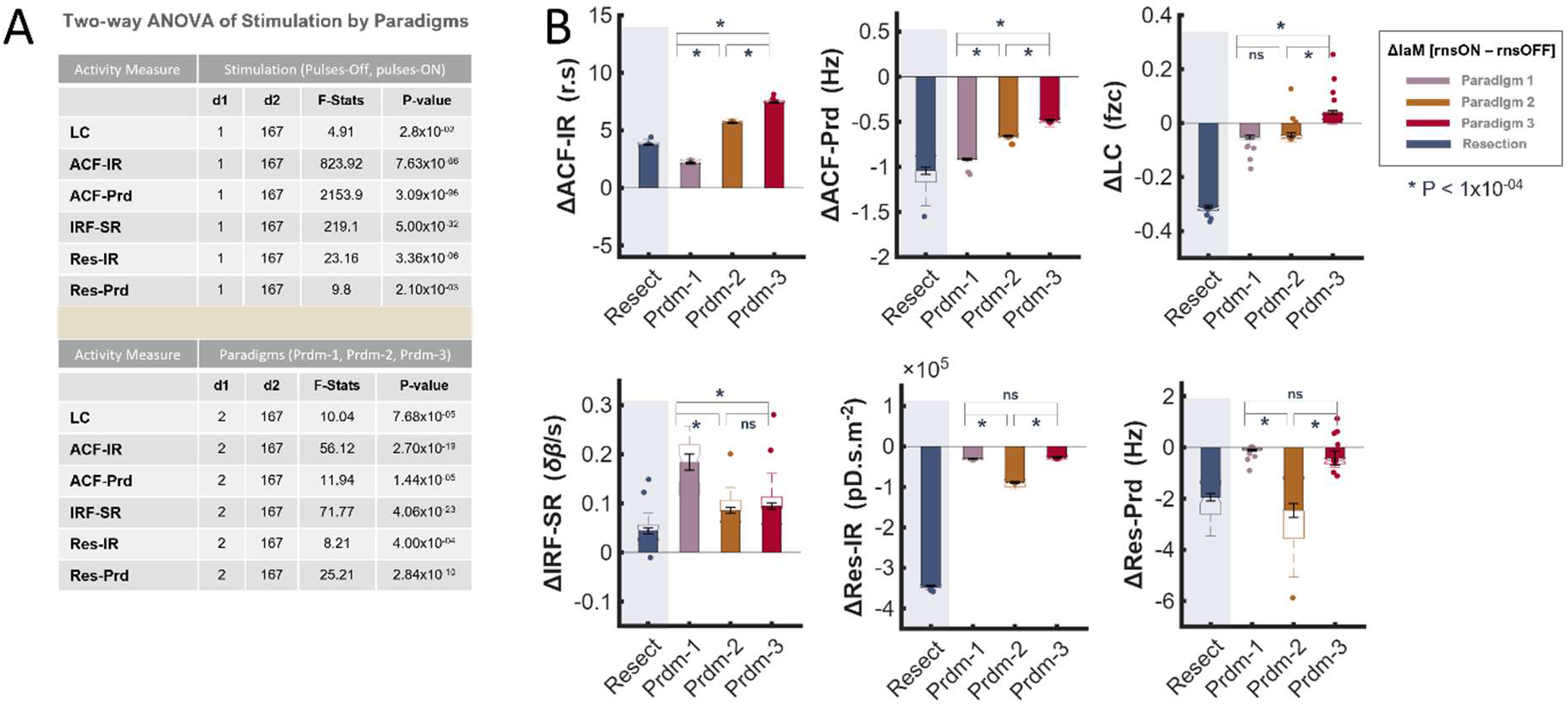
RNS- and resection-evoked changes in interictal activity markers (IaMs). **A)** Summary table of two-way analysis of variance (ANOVA) results testing the effects of stimulation condition (pulses-OFF versus pulses-ON) and RNS paradigms (Paradigm 1, Paradigm 2 and Paradigm 3) on each IaM. Significant main effects of stimulation condition are shown in the top half of the table, and significant effects of RNS paradigm in the bottom half. **B)** Mean net changes in IaMs (ΔIaM) following RNS stimulation and surgical resection. For RNS, ΔIaM represents the change from pulses-OFF to pulses-ON for each stimulation paradigm (Prdm-1, Prdm-2, and Prdm-3, pink, orange, and red bars, respectively). For resection, ΔIaM represents the change from pre-resection to post-resection activity in the PRZ cortex (Resect, blue bars). Net changes were calculated by averaging ΔIaMs across all 28 dR-size configurations. Significant differences among RNS paradigms were observed in the magnitude of ΔACF-IR, ΔACF-Prd ΔIRF-SR, ΔRes-IR, and ΔRes-Prd (p < 0.0001), as well as in both the magnitude and direction of ΔLC markers (p < 0.0001, for IaMs).

Next, to compare the mean net-changes across paradigms, we performed a one-way ANOVA on the ΔIaMs, calculated as the difference between the IaMs derived during pulses-ON and pulses-OFF, separately for each activity marker. Significant differences were observed across paradigms for all activity markers [LC: [F (2, 81) = 37.12, ACF-IR: F (2, 81) = 6.88x10^2^, ACF-Prd: F (2, 81) = 1.99x10^3^, IRF-SR: F (2, 81) = 21.73, Res-IR: F (2, 81) = 6.67x10^2^, and Res-Prd: F (2, 81) = 58.85, p < 1.x10^-05^, for all IaMs]. Post hoc analysis confirmed that all three paradigms consistently evoked either a net increase (ACF-IR, IRF-SR, Fig. 7B, column 1) or a net decrease (Res-IR, ACF-Prd, Fig. 7B, column 2) in activity markers during stimulation. These directional changes were consistent with those observed at the optimum TdRs and were preserved across dR sizes.

Notably, comparisons among the stimulation paradigms revealed significant differences in the magnitude of the mean changes in ACF-IR and ACF-Prd (ΔACF-IR: 2.15 ± 0.077, 5.64 ± 0.052, and 7.42 ± 0.047 r.s, mean ± s.e.m., for Paradigm 1, Paradigm 2 and Paradigm 3, respectively; ΔACF-Prd: -0.91 ± 0.009, -0.66 ± 0.007, and -0.48 ± 0.009 Hz, mean ± s.e.m., Paradigm 1, Paradigm 2 and Paradigm 3, respectively, p < 1.0x10^-05^ for both IaMs) (Fig. 7B, top columns 1, 2). Significant differences in the *magnitude* of the mean IRF-SR change were also observed among paradigms, except between Paradigms 2 and 3 (ΔIRF-SR: 0.18 ± 0.016, 0.08 ± 0.006, and 0.094 ± 0.010 δB/s; mean ± s.e.m., for Paradigm 1, Paradigm 2 and Paradigm 3, respectively, p < 1.0x10^-05^, Fig. 7B, Column 1 - bottom). Likewise, significant differences in the *magnitude* of the mean changes in Res-IR and Res-Prd were observed among paradigms, except between Paradigms 1 and 3 for both markers (ΔRes-IR: -2.99x10^4^ ± 6.72x10^2^, -8.88x10^4^ ± 2.02x10^3^, and -2.69x10^4^ ± 9.71x10^2^ pD.s.m^-2^, for Paradigm 1, Paradigm 2 and Paradigm 3, respectively; ΔRes-Prd: -0.11 ± 0.036, -2.46 ± 0.272, and -0.42 ± 0.090 Hz, Paradigm 1, Paradigm 2 and Paradigm 3, respectively, p < 1.7x10^-06^ for both IaMs) (Fig. 7B, bottom, column 2 and 3). In contrast, LC exhibited paradigm-dependent differences in both the *magnitude* and *direction* of the net change, although no significant difference was observed between Paradigms 1 and 2. Specifically, changes in LC (i.e., ΔLC) values were -0.052 ± 0.0067, -0.043 ± 0.0074, and 0.040 ± 0.010 Fzc, mean ± s.e.m. for Paradigm 1, Paradigm 2 and Paradigm 3, respectively (p < 1.0x10^-05^ for IaMs, Fig. 7B, column 3, top). The detection of these paradigm-specific differences demonstrates the sensitivity of the fUSI-derived IaMs to distinguish RNS stimulation paradigms.

Finally, comparison of RNS- and resection-induced changes revealed similar directional effects across most IaMs. Specifically, the net changes in the IaMs in PRZ after resection of SOZs, averaged across all 28 dR-size configurations were ΔLC: -0.31 ± 0.005 Fzc mean ± s.e.m., ΔACF-IR: 3.79 ± 0.048 r.s, ΔACF-Prd: -1.04 ± 0.040 Hz, ΔIRF-SR: 0.044 ± 0.0063 δB/s, ΔRes-IR: -3.44 ± 0.00016 pD.s.m^-2^, and ΔRes-Prd: -1.95 ± 0.147 Hz, (Fig. 7B, blue bars). Notably, both SOZ resection (Resect, blue bars) and RNS across all paradigms (Prdm-1, -2 and -3, pink, orange, and red bars, respectively), increased ACF-IR and IRF-SR while decreasing ACF-Prd, Res-IR, and Res-Prd. The direction of the LC change following resection was consistent with that observed during RNS Paradigms 1 and 2, but not Paradigm 3. Thus, despite representing distinct therapeutic interventions, SOZ resection and RNS produced similar directional changes across five of the six IaMs.

## Discussion

### General

In this study, we demonstrate that interictal hemodynamic dynamics measured with intraoperative functional ultrasound imaging (fUSI) contain distinct signatures that differentiate seizure-onset zone (SOZ) from surrounding peri-resection cortex (PRZ) and are systematically modulated by both SOZ resection and responsive neurostimulation (RNS). We introduce six ultrafast power Doppler-based interictal activity markers (IaMs) that capture complementary spatial and temporal properties of cortical hemodynamic dynamics. These markers differentiate SOZ from PRZ during interictal periods, reveal changes in surrounding cortical tissue following SOZ resection, and detect robust, paradigm-dependent responses to RNS. Notably, RNS and surgical resection produce concordant directional changes across most of the IaMs, despite representing fundamentally different therapeutic interventions.

These findings unlike previous applications of fUSI in epilepsy ^30–33^, which have primarily focused on hemodynamic changes associated with ictal events, demonstrate that clinically relevant differences in cortical dynamics can be detected during interictal periods. This capability is particularly relevant for intraoperative applications, where spontaneous seizures may not occur during the available imaging period. Current localization of epileptogenic tissue relies predominantly on electrophysiological approaches, including intracranial recordings such as sEEG and iopECoG. Our findings suggest that fUSI could complement these approaches by providing spatially resolved information about interictal cortical dynamics without requiring an ictal event during imaging. Furthermore, the modulation of IaMs following both resection and RNS suggests that fUSI may provide a means not only to characterize epileptogenic tissue but also to assess the immediate effects of therapeutic interventions during epilepsy surgery.

### Interictal neurovascular signatures of seizure-onset zone (SOZ)

Although prior studies have demonstrated abnormal interictal hemodynamic responses associated with SOZ ^34–36^, they have not established consistent, quantitative markers that reliably distinguish SOZ from PRZ during continuous interictal activity. One of the main findings of the current study is that the SOZ exhibits distinct interictal neurovascular dynamics compared with PRZ. Using power Doppler-based interictal activity markers (IaMs), we identified consistent differences between SOZ and surrounding PRZ cortex in properties related to signal energy, local synchrony, and temporal stability. Signal energy describes the magnitude and persistence of fluctuations in the pD signal, whereas local synchrony reflects the degree to which pD signals fluctuate together across neighboring cortical regions. Temporal stability captures a different aspect of dynamics, describing how the pD signal evolves over time, including its settling behavior, persistence, and periodicity. Together, these complementary properties provide a framework for interpreting the distinct interictal neurovascular dynamics observed within the SOZ.

Differences in signal energy were reflected primarily by ACF-IR and Res-IR, both of which were elevated within SOZs relative to PRZs. These findings indicate greater magnitude and persistence of fluctuations in the pD residuals and their corresponding autocorrelation functions within SOZs. In power Doppler imaging, greater signal intensity is associated with greater blood volume within the imaged tissue and, under stable acquisition conditions, can reflect increased local perfusion. Thus, the elevated signal energy observed in SOZs is consistent with increased local hemodynamic activity within epileptogenic cortex. Through neurovascular coupling, increased cerebral blood flow and blood volume can accompany increased neural activity and metabolic demand ^37,38^, providing a potential physiological basis for the greater pD signal energy observed within SOZ. However, because ACF-IR and Res-IR characterize the temporal properties of the pD signal rather than directly measuring blood flow, neural activity, or metabolism, this interpretation should be considered an indirect neurovascular correlate of the epileptogenic state.

Furthermore, the stronger LC observed in SOZs indicates increased linear correlation among local pD signals relative to PRZs. LC quantifies the correlation structure of the hemodynamic signals within the imaged cortical region and therefore provides a computationally tractable measure of local coordination in interictal pD dynamics. Importantly, this measure should not be interpreted as a direct measure of neuronal synchronization or anatomical connectivity. Nevertheless, the higher LC observed in SOZs is consistent with extensive electrophysiological evidence demonstrating increased synchronization and abnormal network coupling in epileptogenic cortex during both ictal and interictal states ^39–41^. Although the relationship between electrophysiological synchronization and correlated hemodynamic fluctuations is complex, our findings suggest that altered local coordination within epileptogenic tissue is also expressed in the interictal neurovascular signal. In contrast to large-scale or long-range functional connectivity metrics, which integrate signals across distributed networks and can be sensitive to spatial-scale differences ^42,43^, LC focuses on correlations among pD signals within the local imaged cortical region. Its consistent elevation in SOZ relative to PRZ across recordings supports LC as a robust marker for distinguishing these regions during interictal activity.

Beyond differences in signal energy and local synchrony, these regional distinctions also have important implications for the temporal dynamics of the underlying pD signals. In this context, temporal stability describes how the modeled hemodynamic dynamics respond to and settle following a transient perturbation, providing information about the persistence of its influence over time. Specifically, the lower IRF-SR observed in SOZ was associated with ARIMA-based models exhibiting more rapidly decaying impulse responses to a unit (i.e., Dirac delta) input, indicating faster settling of the modeled dynamics. This behavior suggests a more tightly constrained temporal regime, in which the influence of a transient input dissipates quickly. In contrast, the higher IRF-SR observed in PRZ was associated with more slowly decaying impulse responses of the fitted ARIMA models, indicating greater temporal persistence of the modeled response following a transient input. Thus, SOZ and PRZ differed not only in signal energy and local coordination, but also in the temporal characteristics of the interictal hemodynamics.

### Resection-induced changes in peri-resection zone (PRZ)

An important finding of this study is that SOZ resection significantly altered interictal dynamics within adjacent PRZ cortex. Prior to resection, these regions exhibited intermediate dynamics, showing elevated local connectivity, residual energy, and signal periodicity, together with lower IRF-SR values compared with post-resection PRZ, but less pronounced than those observed in the SOZ. Thus, before resection, PRZ cortex exhibited an intermediate interictal hemodynamic profile between the SOZ and its post-resection state.

This intermediate profile may reflect the influence of the nearby SOZ on surrounding cortical dynamics. Interictal epileptiform activity originating within SOZ can propagate through functionally connected brain regions, recruiting neighboring cortex in stereotyped spatiotemporal patterns and contributing to an epileptic network that extends beyond the primary seizure focus ^43–47^. Within this framework, the elevated local synchrony and signal energy observed in adjacent PRZ cortex before SOZ resection may reflect the influence of pathological activity originating within the SOZ on surrounding cortical dynamics.

Following resection, we observed consistent reductions in LC, Res-IR, Res-Prd, and ACF-Prd, accompanied by increases in IRF-SR and ACF-IR. Together, these changes indicate a reorganization of multiple aspects of interictal hemodynamic dynamics within the surrounding PRZ, including local synchrony, signal energy, periodicity, and temporal response characteristics. Importantly, because these changes occurred in cortical tissue that remained after resection, they demonstrate that the effects of SOZ removal extend beyond the resection margin.

These findings have important clinical implications. They suggest that fUSI may provide an intraoperative marker of changes in surrounding cortical dynamics following SOZ resection, offering a potential means to assess the immediate network-level effects of surgical intervention. Notably, all patients in this cohort remained seizure-free following resection, suggesting that the observed changes in PRZ dynamics accompanied successful seizure control.

### Novel avenues for improving neurostimulation therapeutics for drug-resistant epilepsy

Neuromodulation provides an important therapeutic option for patients with drug-resistant epilepsy who are not candidates for surgical resection. Among these approaches, RNS is particularly suited for targeting identified epileptogenic regions because stimulation is delivered in response to detected abnormal electrophysiological activity ^20,23,48^. However, the effects of RNS on the local and distributed dynamics of the targeted brain tissue remain incompletely understood ^2520,22^. This knowledge gap is particularly relevant because stimulation parameters can be adjusted across multiple dimensions, yet there are limited approaches for directly assessing their immediate effects on the targeted cortical network.

In this context, combining fUSI with RNS provides an opportunity to characterize stimulation-induced changes in cortical hemodynamic dynamics with high spatiotemporal resolution. In this study, we captured RNS-evoked changes in cerebral hemodynamics and interictal dynamics across three stimulation paradigms. Notably, RNS-induced modulation of interictal activity showed striking directional similarities to the changes observed following surgical resection of SOZ, despite these interventions operating through fundamentally different mechanisms. Specifically, both RNS stimulation and surgical resection were associated with increases in ACF-IR and IRF-SR, alongside decreases in ACF-Prd and Res-IR. Res-Prd also decreased following resection and across all three RNS paradigms, resulting in concordant directional changes across five of the six IaMs.

LC was the only marker for which the direction of the RNS-induced change was not consistent with resection across all stimulation paradigms. LC decreased during Paradigms 1 and 2, consistent with the reduction observed following resection, but increased during Paradigm 3. In addition, both LC and Res-Prd exhibited strong dR-dependent behavior, with the magnitude and, for LC, direction of stimulation-induced changes varying across spatial discretization scales and stimulation paradigms. These findings indicate that the effects captured by these markers are particularly sensitive to both stimulation parameters and the spatial scale at which interictal dynamics are evaluated.

Overall, although further work is required to establish the clinical implications of these findings, they demonstrate the potential of fUSI as a tool for monitoring the effects of neurostimulation on cortical dynamics. The ability to distinguish responses across RNS stimulation paradigms further suggests that fUSI could ultimately provide a functional readout for evaluating and optimizing stimulation parameters in individual patients.

### Limitations and future directions

While we robustly detected epilepsy-related alterations in interictal activity within the SOZ, as well as strong evoked responses to both surgical resection and RNS, it is important to acknowledge that the anatomical classification of SOZ and PRZ in this study was based on the clinical localization established during standard Phase I and Phase II epilepsy evaluations, including electrophysiological recordings with ECoG and/or sEEG. Although these approaches provide the clinical basis for identifying epileptogenic tissue and guiding surgical treatment, their spatial sampling is inherently constrained by electrode coverage and placement. Consequently, epileptogenic activity occurring outside the sampled regions or distributed across broader networks may not be fully captured. Importantly, all patients who underwent SOZ resection remained seizure-free during the available postoperative follow-up, providing clinical support that the regions identified and resected encompassed the tissue necessary for seizure generation in these patients. Within this clinically validated framework, the consistent differences observed between SOZ and PRZ demonstrate that fUSI-derived IaMs capture distinct interictal hemodynamic dynamics associated with these regions.

Although the enhanced sensitivity of fUSI enables high-resolution imaging of cerebral hemodynamics, its application is currently limited by the attenuation and distortion of high-frequency acoustic waves by the adult skull ^49–51^. As a result, most preclinical and clinical fUSI applications have been minimally invasive, except in young mice with thin skulls (8-12 weeks old) ^52^ and in pediatric participants, where the fontanelle provides a natural acoustic window ^29,33^. Consequently, transcranial fUSI acquisition in awake adults remains technically challenging. Recent technological advances, however, suggest that these limitations may be increasingly addressed ^53^. For instance, we previously demonstrated fUSI recordings of task-related brain activity in an awake adult through a permanently implanted polymeric cranial acoustic window ^53^. More recently, modulation of skull acoustic properties using a topically applied chelating agent enabled transcranial fUSI through the intact skull in mice, while experiments using ex vivo human skull demonstrated the potential translational applicability of this approach ^54^. In parallel, intravenously administered microbubble contrast agents have enabled transcranial measurements of cerebral hemodynamics^55,56^. Together, these developments demonstrate multiple emerging strategies for overcoming the skull barrier and support the feasibility of repeated, minimally invasive or non-invasive fUSI in human brain studies. These advances may expand the use of fUSI for investigating pharmacological effects, neurosurgical interventions, and neurostimulation strategies in awake patients with epilepsy during interictal periods. In the present study, neither implanted acoustic windows nor contrast agents were required, because fUSI was performed during standard-of-care craniotomy for SOZ resection or RNS implantation, which provided direct acoustic access to the cortical surface.

Furthermore, although anesthetic agents can influence cerebral blood flow and thus affect pD signals, we mitigated this concern by maintaining a relatively stable level of anesthesia throughout the surgical procedures. For the RNS experiments, pD changes were quantified relative to baseline recordings (i.e., stimulation OFF), minimizing anesthesia-related differences between stimulation conditions.

Additionally, we calculated %pD changes relative to baseline recordings obtained immediately prior to the RNS stimulation, thereby mitigating potential confounds of anesthesia across conditions. Future fUSI studies in awake patients will further extend our characterization of SOZ- and PRZ-specific interictal dynamics and RNS-evoked responses under natural physiological conditions.

## Materials and Methods

### Patients and surgical procedures

A total of nine (N = 9) patients (3 females and 3 males; age range, 27 – 51 years) were recruited for intraoperative fUSI signal of the brain during seizure-onset zone resection and responsive neurostimulation (RNS) device implantation. Two patient cohorts were included. The first group (N=6) consisted of individuals with drug-resistant focal epilepsy who underwent standard-of-care resection of seizure-onset zones under general anesthesia. The second cohort (N=3) also had severe drug-resistant epilepsy but received standard-of-care implantation of an FDA approved responsive neurostimulation device (RNS; Neuropace neurostimulator Model RNS 320 ® system) under general anesthesia. All surgeries were conducted at the Keck School of Medicine of the University of Southern California. Intraoperative fUSI was performed by placing a miniaturized 15.6-MHz linear array probe through a cranial window onto the cortical surface (Fig. 1 B-C) to record hemodynamic activity. Informed consent was obtained from all patients with risk and study procedures clearly explained, in accordance with protocols approved by the USC Institutional Review Board (HS-19-00994).

### Functional ultrasound imaging data acquisition

A commercial functional ultrasound acquisition system, Iconeus One (Iconeus, Paris, France) was used to record brain activity via a linear array probe inserted through a cranial window (Fig. 1 B-C). The probe had 128 elements, a 15.6 MHz center frequency and 0.1 mm pitch, generating pD images with an in-plane spatial resolution of 100 μm × 100 μm, slice thickness of 400 μm, and a FOV of 12.8 mm (width) × 9.1 mm (depth). The probe was mounted on a surgical articulating arm and fixed steadily throughout the experiments with the FOV transverse to the cortical surface of the brain (Fig. 1 B, C). To increase the signal-to-noise-ratio, each image was generated from 200 compounded frames acquired at 500 Hz frame rate, using 11 tilted plane waves separated by 2° (i.e., from -10° to +10° increment by 2°). The pulse repetition frequency was 5500 Hz. Imaging was performed using a real-time continuous acquisition of successive blocks of 400 ms (with 600 ms pause between pulses) of compounded plane wave imaging – resulting in a final beamformed composite pD image at a rate of 1 Hz. The acoustic amplitudes and intensities of the fUSI sequence remained below the Food and Drug Administration limits for ultrasonic diagnostic imaging (FDA, 510 (k)), yielding a mechanical index of 0.44. The Intensity Spatial peak temporal average (ISPTA) was 120 mw/cm^2^, with a maximum temperature rise limit of +1.2°C at the lens contact.

### Data processing

A built-in phase-correlation based sub-pixel motion registration ^57^ and singular-value-decomposition based clutter filtering algorithms ^58–60^, included in the Iconeus One acquisition system software were used to separate tissue motion signal from blood signal to generate relative pD signal intensity images ^27^. We employed rigid motion correction techniques ^61^ that have successfully been used in fUSI ^50,62,63^ and other neuroimaging studies ^61,64^, to remove potential physiological and motion artifacts. This was combined with in-house high frequency smoothing filtering, utilizing a lowpass filter with normalized passband frequency of 0.04 Hz, with a stopband attenuation of 60 dB that compensates for delay introduced by the filter, to remove high-frequency fluctuations in the pD signals.

### Preoperative ictal and intraoperative interictal activity mapping

To localize seizure-onset zones (SOZs), patients first underwent extensive clinical, imaging and electrophysiological assessments to plan invasive EEG studies. The seizure-onset zone was then defined as the first region displaying seizure during pre-operative Phases I and II interictal and seizure onsets stereo-encephalography (sEEG) assessments, as per standard of care. Additional intraoperative electrocorticography (iopECoG) complementary mapping assessments were conducted pre- and post-resection of the SOZs (Figure 1B). Depth electrodes and/or strip grids were placed over both SOZ (Fig. 1C-a) and adjacent presumed peri-resection zone cortex outside of the planned resection (peri-resection zone, PRZ) (Fig. 1C-c). After resection, iopECoG recordings were also obtained to evaluate the surgical removal (Fig. 1C-e), allowing comparison of interictal activity before and after resection of the seizure zones (Fig. 1B, C). Interictal fUSI recordings were acquired from SOZs as well as adjacent PRZs. Recordings were obtained both pre-resection (Fig. 1C-b, d), and post-resection (Fig. 1C-f), from the frontal (N=2) and temporal (N=4) lobes across six patients: P1 (left temporal), P2 (left frontal), P3, P4, P5 (right temporal), and P6 (left frontal / frontal cortical dysplasia) (Fig. 1B).

### Markers of cortical interictal activity

The analysis of fUSI interictal activity to determine functional differences between SOZ and PRZ presents unique challenges not typically encountered in conventional functional neuroimaging. Standard neuroimaging studies generally rely on baseline reference recordings against which experimental manipulations (e.g., pharmacological administration, sensory or motor task, audiovisual stimuli, electromagnetic stimulation) are compared. In contrast, no such baseline exists in the presence study – there is no external intervention beyond the intrinsic effects of epilepsy on seizure-onset zones. Another challenge arises from the nature of pD signal, which reflects blood flow in arbitrary units – making it impossible to directly compare activity across recording sessions. To address these limitations, we developed an analytical framework designed to capture intrinsic functional activity and local network properties embedded in the interictal pD signal. To do so, for each 3D spatiotemporal pD dataset (*space* × *time*) recorded from SOZs and adjacent PRZs, pre- and post-resection, the FOV was divided into non-overlapping rectangular discretized regions (dR) of size [*m* × *n, column* × *rows*]. The pD intensity signals were then averaged across all pixels within each dR, yielding *m × n* total discretized regions (TdR) time-series signals (Fig. 2A, B). The signal, *x_i_* (*t*), in each dR thus represents an indirect measure of local brain activity coupled to blood flow dynamics. Treating each dR as a local activity source, we derived six interictal activity-markers (IaMs) to quantify functional activity and connectivity properties: i) Local connectivity (LC) ii) autocorrelation function impulse response (ACF-IR), iii) autocorrelation function periodicity (ACF-Prd), iv) impulse response function settling rate (IRF-SR), v) pD residuals impulse response (Res-IR), vi) and pD residuals periodicity (Res-Prd) (Fig. 2A). Refer below for details of each interictal activity measure. To evaluate how dR size influenced these markers, we tested 28 dR-size configurations ranging from 3 × 2 to 16 × 16 (6-256 TdRs), corresponding to dR areas of 1.94-0.046 mm^2^ (Fig. 2B). This analysis allowed us to identify the dR size that best captures local network activity for each IaM and to determine the configuration yielding the largest mean difference when comparing interictal signals across conditions – i.e., pre-resection seizure-onset zone vs. pre-resection peri-resection zone, pre-resection peri-resection zone vs. post-resection peri-resection zone, and pre-resection seizure-onset zone vs. post-resection peri-resection zone. Finally, to assess the statistical significance of these differences, *Student’s t-Test* were performed on the mean IaMs, averaged across recordings and TdRs, comparing cortical SOZs and adjacent PRZs before and after resection.

### Local connectivity (LC)

The local connectivity indirectly quantifies local neural connectivity or the degree of synchronization between non-overlapping signals acquired from a cortical region within the fUSI FOV. It is assessed using Pearson’s cross-correlation coefficients (CC) computed across non-overlapping discretized regions (dRs) of the fUSI recordings (see above for more details). Each dR provides a mean pD time series, *x_i_* (*t*), that indirectly reflects local neural activity coupled to blood flow dynamics.

To characterize how epilepsy alters local functional connectivity in cortical seizure-onset zones, we first modeled each dR time series using an autoregressive integrated moving average, ARIMA (1,1,1) model ^65^. In this configuration, the autoregressive component AR(1) predicts the current value from the immediately preceding value, the integrated component I(1) applies single differencing to reduce non-stationarity, and the moving average term MA(1) incorporates the previous lag error to refine the prediction. Note that the ARIMA [1,1,1] model was selected to provide stable, generic representations of cortical pD signals within the dRs for subsequent analyses. This approach introduced quasi-stationarity, reduced seasonality, and ensured consistent modeling across interictal conditions. While tailoring ARIMA parameters individually for each dR could, in theory, improve stationarity and seasonality removal, such optimization was impractical given the large number of signals analyzed (ranging from ∼124 to 7,936 per dataset). Moreover, a single standardized model configuration ensured uniformity and computational feasibility, whereas optimizing separately would not allow identification of a universally optimum configuration. Finally, residuals, *ε_i_*(*t*), were obtained by subtracting from the actual pD signals *x_i_* (*t*) their ARIMA model representations 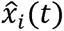 (Eq. 1).

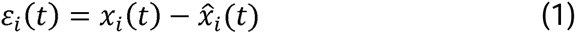

Pearson’s cross-correlation coefficients (CCs) were computed between the resulting pD signal residuals, *ε_i_*(*t*) of the dRs (Fig. 2A). The coefficients across all dR pairs were then averaged to yield the local connectivity measure for each fUSI recording.

### Impulse response function settling rate (IRF-SR)

The impulse response function settling rate (IRF-SR) quantifies how quickly the transfer function of an ARIMA-derived model converges to steady-state when subjected to a unit impulse input (i.e. Dirac delta function input). To assess how epilepsy modulates cortical blood flow dynamics and local neural network synchrony (i.e., degree of matching and interdependent behavior across a given timescale), we hypothesized that the temporal characteristics of ARIMA representations of pD signals depend on the degree of neural connectivity and synchrony. Specifically, because cortical seizure-onset zones typically exhibit increased neuronal population activity (as reflected by stronger sEEG and iopECoG activity compared with normal cortex), we anticipated measurable differences in signal properties – such as synchrony and temporal decay (i.e., time to steady-state after perturbation) – that would be captured by ARIMA modeling. To test this hypothesis, we examined the impulse response dynamics of ARIMA representations of interictal fUSI signals. For each dR signal, we applied a unit impulse input to the ARIMA-derived transfer function and evaluated the rate at which the model response converged to steady state. The settling rate was quantified by fitting a linear model to the final 66.7% of the IRF curve and extracting its slope (i.e., first-order coefficient; Fig. 2A, b, c). To compare activity across cortical regions, we averaged these slopes across dRs and recordings from seizure-onset (pre-SOZ) and peri-resection (pre-PRZ) zones before resection, as well as from peri-resection zones, post-resection (post-PRZ). The resulting IRF-SR values provided a measure of differences in temporal response dynamics across conditions.

### Autocorrelation Function Impulse Response (ACF-IR) and Periodicity (ACF-Prd)

To investigate the local linear dependency and oscillatory properties of cortical interictal activity, we computed autocorrelation functions (ACFs) for each dR pD time-series signal in each fUSI recording. This approach assumes that epilepsy alters both blood flow dynamics and local neural synchrony in cortical SOZs over time. Given the stronger sEEG and iopECoG electrical activity observed in SOZs, we expected to detect distinct dependencies in oscillatory patterns and energy demand, reflected in increased blood flow compared to normal cortical regions. The autocorrelation function assesses the strength (i.e. impulse) and rate of cyclic changes (i.e. periodicity) properties contained in pD signal acquired from SOZ and surrounding PRZ regions. Autocorrelation ***p**_k_* (Eq. 2) quantifies the linear dependence of a time-series with itself at different time lags. It is defined as the ratio of the covariance between the signal and a delayed copy of itself at lag-k to the variance at lag zero ^66–68^.

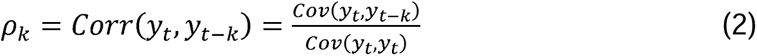

For each dR’s pD time-series, we generated the autocorrelation (*p_k_*) across 50 (RNS pD signal) or 116 (resection pD signal) lags (L), (*p*_O_ … *p_k_*, k = 1, 2, …. L). To assess the strength of temporal linear dependence, we fitted a curve to the autocorrelation function (ACF, i.e., autocorrelation coefficients across lag steps) and calculated the area under this curve, which we defined as the autocorrelation function impulse response (ACF-IR). To capture the oscillatory component, we defined the autocorrelation function periodicity (ACF-Prd) as the frequency at which the lag correlation coefficients of the ACF alternated between peak (maxima) and trough (minima) values, i.e. cycles per lag (Fig. 2A b, c). To determine the largest difference in the ACF-Prd measure when we compare pD signals acquired from seizure-onset and peri-resection zones pre- and post-resection, we determined the optimum ACF coefficient thresholds, for each pair of conditions compared. The thresholds set the limit on the minimum ACF correlation-coefficient value to include in calculating the ACF-Prd measure. Fig. S1G shows tuning curves of mean ACF-Prd differences as a function of correlation coefficient thresholds across conditions: pre-resection SOZ (pre-SOZ) vs. pre-resection PRZ (pre-PRZ), (red-curve), pre-PRZ vs. post-resection PRZ (post-PRZ), (blue curve) and pre-SOZ vs. post-PRZ (green curve). To compare the signals acquired pre-resection, from pre-SOZ and pre-PRZ regions, we utilized ACF correlation-coefficient threshold of 0.1345 as it results in the largest mean difference (red curve). Additionally, a 0.0055 ACF correlation coefficient threshold produced the largest difference in ACF-Prd, when signals acquired from the same cortical PRZs, pre- and post-resection of the SOZs, are compared (pre-PRZ vs. post-PRZ, blue curve) (Fig. S1G). ACF correlation-coefficient threshold of 0.0235 was utilized when comparing signals acquired pre-resection from seizure-onset zones to signals acquired post-resection from peri-resection zones (Fig. S1G, green curve).

### pD Residual Impulse Response (Res-IR) and Periodicity (Res-Prd)

To investigate differences in energy demand, reflected in cerebral blood flow during interictal activity within SOZs versus surrounding PRZs, we derived the pD residual impulse response (Res-IR) for each dR’s pD time-series signal. This approach assumes that epilepsy modulates hemodynamic activity, as suggested by iopECoG differences between SOZs and PRZs. To compute the Res-IR, we first generated residuals, *ε_i_*(*t*) by subtracting the ARIMA model representation of each dR signal, 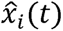, from the actual pD time-series signal, **x_i_**(*t*), in the dR (Eq. 1). The area under this residual curve provided the Res-IR measure. Additionally, we examined unique oscillatory patterns in the residual signals by calculating the residuals periodicity (Res-Prd). Res-Prd was defined as the rate at which the residuals time-series alternated between peak and trough values (i.e. cycles per sec) (Fig. 2A b, c). Note that the Res-IR marker was normalized by dividing the IaM values by the dR area, ensuring that the IaM values were independent of the dR area.

### Responsive neurostimulation protocol and parameters

All patients receiving cortical RNS underwent a standardized stimulation protocol consisting of three standard-of-care paradigms (total duration: 17 min 50 s; Fig. 5B). Continuous fUSI data were acquired through a cranial window throughout the entire stimulation protocol, beginning with a 2-min baseline recording. This was followed by sequential RNS stimulation using Paradigm 1-3, each separated by rest periods. After completion of Paradigm 3, an additional 2-min fUSI recording was acquired during the post-stimulation period (Fig. 5C). RNS Paradigm 1 consisted of five pulses-ON and - OFF cycles, with each pulses-ON period delivering a single 100-ms stimulation pulse to the brain. RNS Paradigm 2 included three pulses-ON and -OFF cycles, with each pulses-ON period consisting of ten 100-ms stimulation pulses, lasting approximately 24 s. RNS Paradigm 3 consisted of five pulses-ON and -OFF cycles, each pulses-ON period delivering five 100-ms stimulation pulses, lasting approximately 11 s (Fig. 5C). Stimulation pulses were delivered with a pulse width of 160 µs, frequency of 200 Hz, charge density of 2 µC/m^2^ and current amplitude 4 mA through alternating bipolar pairs in each electrode (+-+- or -+-+; Fig. 5E). All three patients received identical RNS stimulation protocol using the same device model (Neuropace neurostimulator Model RNS 320 ® system). The RNS strip and/or depth electrodes were implanted primarily in temporal and frontal cortices: in P7, anterior and posterior to focal cortical dysplasia (strip electrodes), P8, within the hippocampal region (strip and depth electrodes), P9, in left anterior insula (depth electrodes) and interhemispheric region (strip electrodes).

### Effects of responsive neurostimulation on brain hemodynamics and interictal activity markers

We generated statistical parametric maps (SPM) to characterize the spatial and temporal changes in interictal brain activity evoked by the three RNS stimulation paradigms. These SPMs visualize brain regions exhibiting significant (p < 0.01, with FDR correction) increases or decreases in CBV when the RNS device was active. To generate the SPMs, we extracted the pD signal acquired during the pulses-ON and pulses-OFF periods and organized them into two separate 3D data matrices two *X* × *Y* × *t* (2D pD image slice × time): one representing the RNS pulses-ON activity and the other representing the pulses-OFF activity. Each matrix was averaged across all trials for each stimulation paradigm. The last 5 s of activity during the pulses-OFF periods served as reference for each paradigm. Brain regions showing significant CBV changes during the pulses-ON periods were identified by performing pixelwise, two-sided *Student’s t-Test* comparing pD signals between pulses-ON and the reference pulses-OFF conditions, with FDR correction (Benjamini-Hochberg^69^) applied for multiple comparisons. An independent SPM was generated separately for each RNS paradigm. For RNS Paradigm 1, individual stimulation pulses lasted 100 ms, a duration shorter than the temporal window required to reliably capture hemodynamic responses. Accordingly, a 5-s window immediately following pulse onset was defined as the pulses-ON period. This interval enabled the pD signal to capture the full hemodynamic response to stimulation while providing sufficient data for comparison with the corresponding pulses-OFF reference activity. To visualize the most strongly modulated regions, we displayed the top 5% of significantly modulated voxels ranked by z-scored %CBV changes relative to the reference period (i.e., the final 5 s of the pulses-OFF periods). This approach enabled clearer visualization of stimulation-induced responses while avoiding overcrowding of the SPM maps. Finally, the activations were overlaid onto a mean pD grayscale vascular map derived from the baseline recordings to visualize the spatial distribution of the significantly modulated regions. To assess the temporal dynamics of the activations, we also generated event-related-average (ERA) waveforms. The ERA waveforms represent the percentage changes (%CBV) over time, calculated from the significantly activated pixels (increase and decrease CBV) relative to the reference activity. To obtain a representative temporal profile of the overall response, we computed a pseudo-global ERA waveform by averaging the activity across the top 5% of significantly activated pixels.

Next, we extracted the pD signals recorded during the pulses-ON and pulses-OFF periods for each of the three RNS paradigms to examine how each RNS stimulation paradigm influenced interictal neurovascular dynamics. For each paradigm, the pulses-ON and pulses-OFF signals were averaged across trials, resulting in two *X* × *Y* × *t* mean 3D data matrices (pD image *slice* × *time*): one for the pulses-OFF and one for the pulses-ON period. Because the durations of the pulses-OFF periods (25 s, 66 s, 29 s for Paradigm 1, Paradigm 2 and Paradigm 3, respectively) were longer than those of the pulses-ON periods (5 s, 24 s, and 11 s, respectively, Fig. 5C), we applied a linear interpolation algorithm [*interpn()* function with “linear” as “method” input, Matlab R2024b] to the pulses-ON signals to normalize the number of sample points. This ensured balanced datasets for subsequent analyses. To assess RNS-evoked modulation of interictal neurovascular dynamics, we computed the six IaMs using the framework described in Fig. 2 and compared their mean values between pulses-OFF and pulses-ON periods. Specifically, the 3D data matrices for each condition were discretized into *m* × *n* non-overlapping rectangular dRs, and the six IaMs (IRF-SR, ACF-IR, ACF-Prd, Res-IR, Res-Prd and LC) were computed. We hypothesized that both the direction and magnitude of RNS-evoked changes in each activity measure may depend on dR size and stimulation paradigm. To examine this spatial dependence, we evaluated 28 dR discretization configurations, corresponding to dR areas ranging from 1.94 to 0.046 mm^2^ (Fig. 2B). To mitigate potential confounding effects of dR size, we additionally quantified the overall effect of RNS by averaging IaM values during pulses-OFF and pulses-ON periods across all 28 dR configurations and subsequently across recordings. Finally, a two-way analysis of variance (ANOVA), followed by post hoc multiple-comparison testing was used to evaluate differences in RNS effects between stimulation levels (pulses-OFF vs. pulses-ON) and RNS paradigms (Paradigm 1, Paradigm 2, and Paradigm 3).

### Software analysis

All data pre- and post-processing and statistical analysis were performed using Matlab Version (R2024b, R2025b).

## Supporting information

KAgyemanEtAl suplementary

## Data availability

The datasets generated and analyzed during the current study are available from the corresponding authors on reasonable requests. The datasets cannot be used for commercial purposes, and we are unable to fulfill requests that would violate patient confidentiality, ethical guidelines, or patient consent agreements.

## Acknowledgments

We thank the participants that made this study possible. This work was supported by “The USC Neurorestoration Center” at the University of Southern California, and the Mann Engineering & Medicine Pilot Grant Program.

## Author Contributions

K.A.A., D.J.L., C.L. and V.N.C conceived the study.; D.J.L., B.L. and C.L. performed the surgeries; D.J.L., K.A.A, W.C., and K.W. acquired the functional ultrasound data; K.A.A. performed the functional ultrasound data processing, and developed the interictal data markers analysis pipelines; K.A.A. and V.N.C drafted the manuscript with contributions from J.R., D.J.L., R.J.M and C.L.; R.J.M provides clinical outcomes of the cases. All authors edited and approved the final version of the manuscript; C.L. and V.N.C. supervised the research.

