## Supplementary material for "Interictal functional ultrasound imaging in human epilepsy resection and responsive neurostimulation surgery": KAgyemanEtAl suplementary

### Supplementary Materials

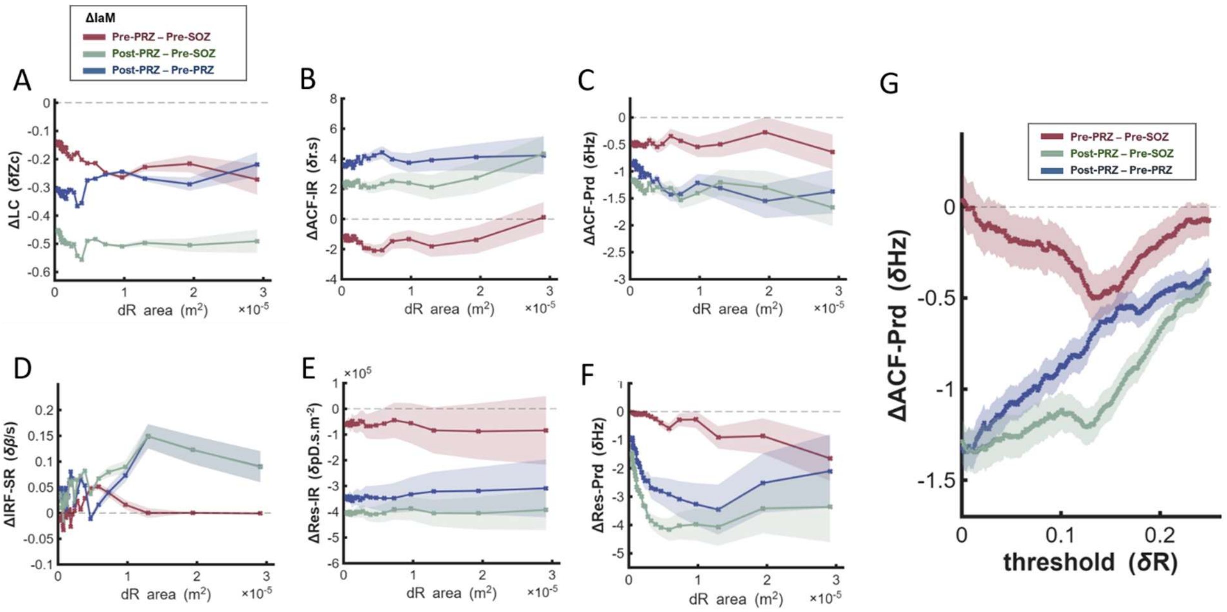


**Figure S1. Spatial-scale dependence of interictal activity markers (IaMs) and ACF-Prd threshold tuning. A)** Changes in the local connectivity (LC) as a function of discretized region (dR) size for comparisons between brain conditions: Pre-PRZ vs. Pre-SOZ (red curve), Post-PRZ vs. Pre-SOZ (green curve) and Post-PRZ vs. Pre-PRZ (blue curve). Shaded regions denote the standard error of the mean across dRs and recordings. Twenty-eight dR configurations ($3 \times 2$ up to $16 \times 16$, corresponding to 6-256 total discretized regions and areas of 1.94-0.046 $mm^2$) were evaluated to identify the dR size yielding the optimum (i.e., largest) mean differences in each IaM. **B)** Same as (A) for the pD residual impulse response (Res-IR) measure. **C)** Same as (A) for the pD residual periodicity (Res-Prd) measure. **D)** Same as (A) for the impulse response function settling rate (IRF-SR) measure. **E)** Same as (A) for the autocorrelation function impulse response (ACF-IR) measure. **F)** Same as (A) for the autocorrelation function periodicity (ACF-Prd) measure. **G)** Tuning curves of mean ACF-Prd differences as a function of correlation coefficient thresholds for comparisons between Pre-PRZ vs. Pre-SOZ (red curve), Post-PRZ vs. Pre-SOZ (green curve) and Post-PRZ vs. Pre-PRZ (blue curve). The shaded regions denote the standard error of the mean across TdRs and recordings.

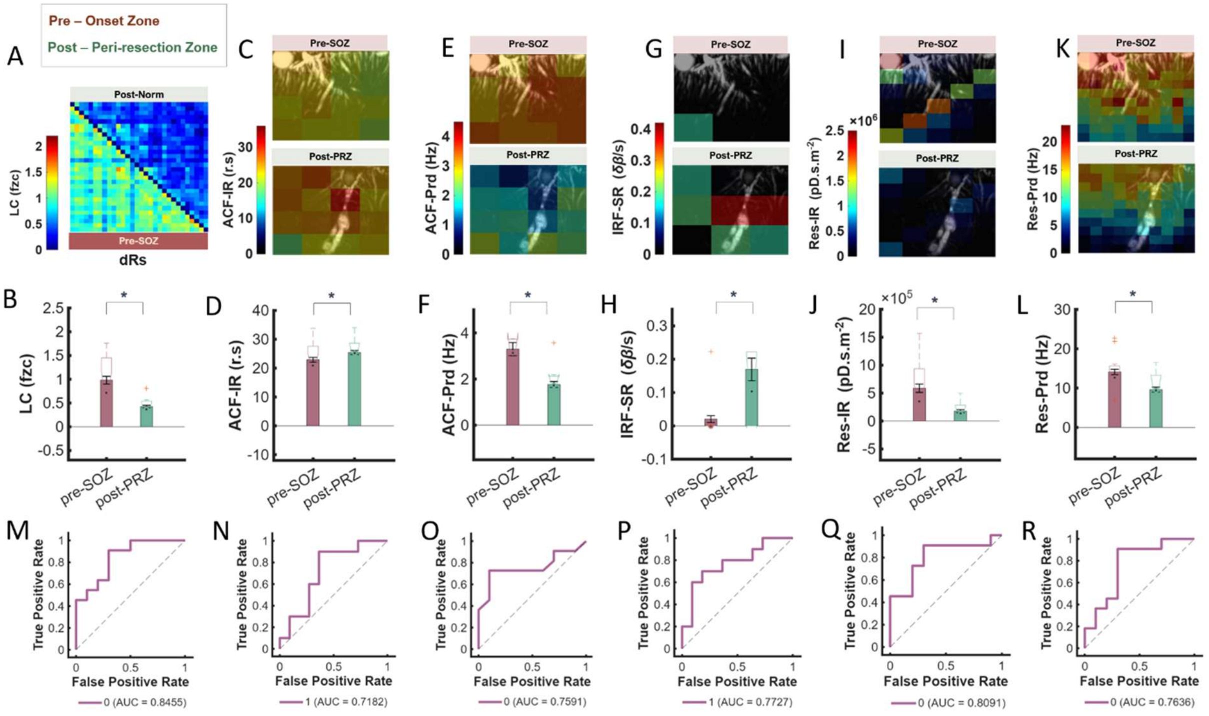


**Figure S2. Interictal activity markers (IaMs) in pre-resection cortical seizure-onset (pre-SOZ) and post-resection peri-resection (post-PRZ) zones. A)** Mean Pearson cross-correlation coefficient matrix computed from pD residuals at the optimum TdR (30) for the LC measure and averaged across recordings. LC of seizure-onset zone is shown in the bottom-left triangle (red) and peri-resection zone, post-resection in the top-right triangle (green). **B)** Mean local connectivity derived from Pearson cross-correlation coefficients, showing higher values in pre-SOZ (red) relative to post-PRZ, (green) after resection ($p < 1.0\times10^{-6}$). **C)** Mean autocorrelation function impulse response (ACF-IR) for each dR in pre-SOZ, (top panel) and post-PRZ (bottom panel), averaged across recordings at the optimum discretization (TdR = 16) and overlaid on representative grayscale, mean pD vascular maps. ACF-IR represents the area under the ACF waveforms obtained by fitting the autocorrelation function computed over 116 lag steps. **D)** Mean ACF-IR across dRs and recordings, showing a significant difference between pre-SOZ and post-PRZ ($p < 1.0\times10^{-2}$). **E)** Mean autocorrelation function periodicity (ACF-Prd) for each dR in pre-SOZ (top) and post-PRZ (bottom), averaged across recordings at the optimum discretization (TdR = 16) and overlaid on representative grayscale, mean pD vascular maps. ACF-Prd represents cycles per 1s lag step derived from peak-to-trough oscillations in dR ACF waveforms over 116 lag steps. **F)** Mean ACF-Prd across dRs and recordings, showing significant difference between pre-SOZ (red) and post-PRZ (green) (p <

$1.0x10^{-6}$). **G)** Mean impulse response function settling rate (IRF-SR) for each dR in pre-SOZ (top) and post-PRZ (bottom), averaged across recordings at the optimum discretization (TdR = 30) and overlaid on representative grayscale, mean pD vascular maps. IRF-SR represents the slope derived from dR IRF waveforms evaluated up to 1000 s after perturbation. **H**) Mean IRF-SR across dRs and recordings, showing lower values in pre-SOZ (red) than post-PRZ (green) ($p < 0.05$). **I)** Mean pD residual impulse response (Res-IR) for each dR in pre-SOZ (top) and post-PRZ (bottom), averaged across recordings at the optimum discretization (TdR = 100) and overlaid on representative grayscale, mean pD vascular maps. **J)** Mean Res-IR across dRs and recordings, showing significant difference between pre-SOZ (red) and post-PRZ ($p < 1.0x10^{-5}$). **K)** Mean pD residual periodicity (Res-Prd) for each dR in pre-SOZ (top) and post-PRZ (bottom), averaged across recordings at the optimum discretization (TdR = 30) and overlaid on representative grayscale, mean pD vascular maps. **L)** Mean Res-Prd across dRs and recordings, showing lower values in post-PRZ (green) than pre-SOZ ($p < 1.0x10^{-5}$). **M)** Receiver operating characteristics (ROC) curve and corresponding area under curve value (AUC) for distinguishing pre-SOZ and post-PRZ recordings using LC at the optimum TdR. **N)** Same as M for ACF-IR measure. **O)** Same as M for ACF-Prd measure. **P)** Same as M for IRF-SR measure. **Q)** Same as M for Res-IR measure. **R)** Same as M for Res-Prd measure.

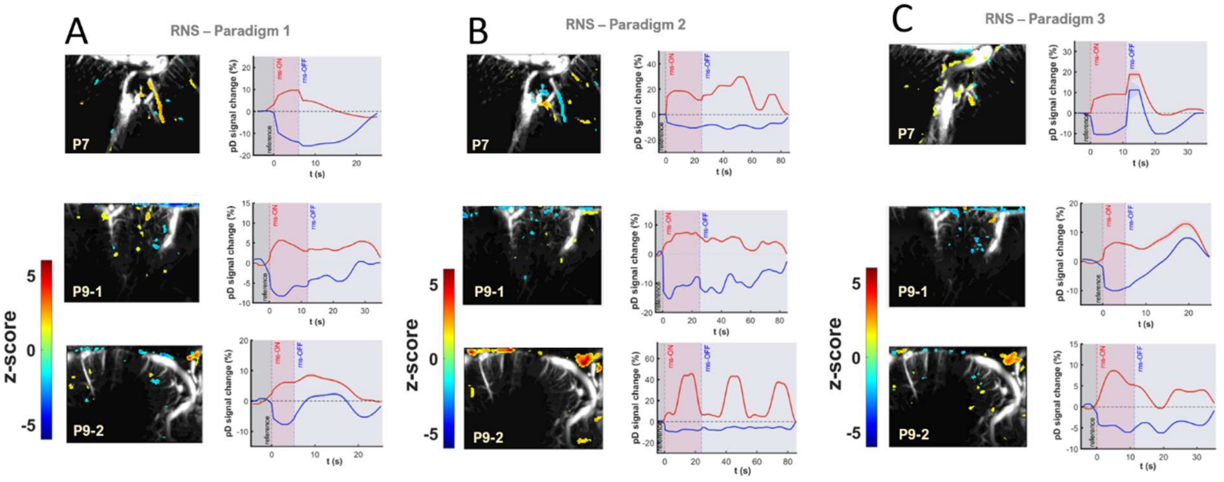


**Figure S3. Statistical parametric maps (SPM) and event-related average (ERA) waveforms of RNS-evoked hemodynamic responses. A) (left panel)** SPMs for Patients 7 and 9 (P7, P9-1, P9-2) showing localized areas with significantly increased and decreased CBV relative to reference activity (last 5 s of pulses-OFF periods; two-sided *Student's t-test*, $p < 0.01$,

FDR corrected across pixels) for RNS Paradigm 1. SPMs are overlaid on grayscale, mean baseline pD images, and the top 5% of significantly activated pixels (based on %CBV changes z-scores) are displayed. **A) (right panel)** ERA waveforms of the activated pixels in the SPMs that exhibit strong CBV increases (red curve) or decreases (blue curve), during the RNS pulses-ON periods for P7, P9-1, P9-2, using RNS Paradigm 1. The pseudo-global ERA waveforms represent the mean activity across the top 5% activated pixels. **B)** Same as panel **(**A) for RNS paradigm 2. **C)** Same as panel (A) for RNS paradigm 3.

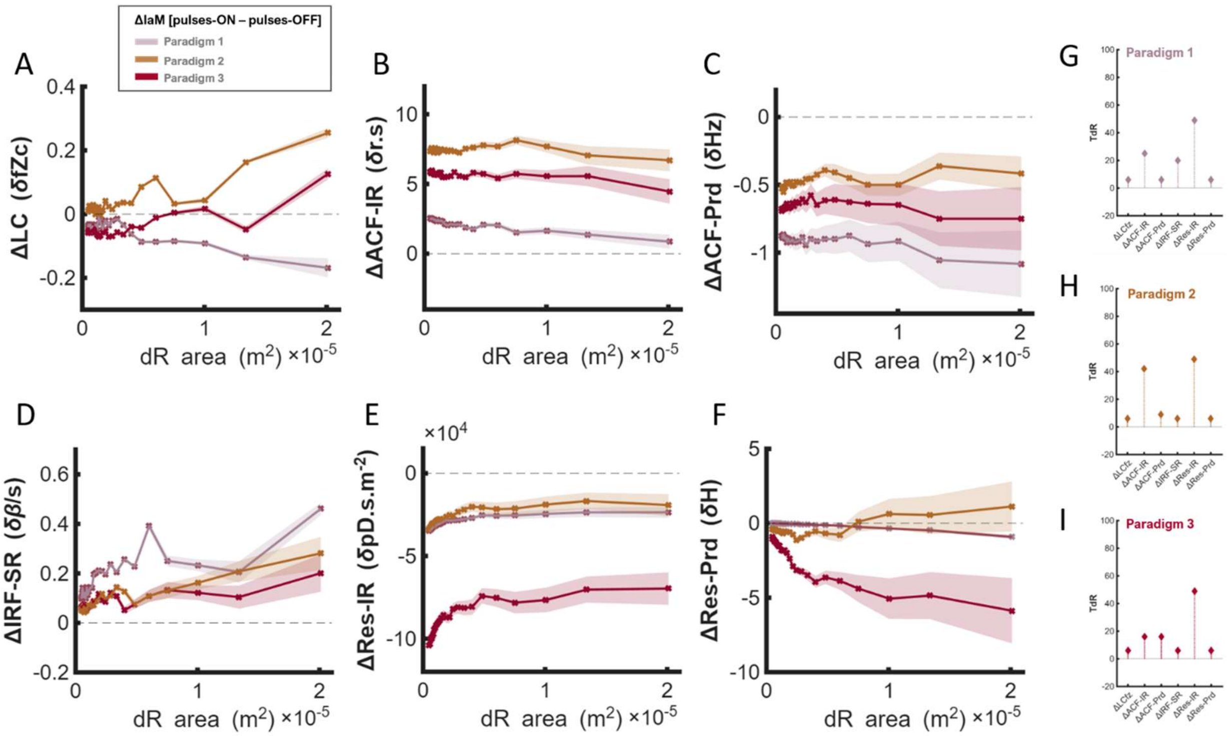


**Figure S4. Spatial-scale dependence of RNS paradigm-induced changes in interictal activity markers (IaMs). A)** Changes in local connectivity (LC) during RNS pulses-ON periods with respect to reference activity during the pulses-OFF periods as a function of dR size for RNS Paradigms 1-3 (pink, orange and red curves, respectively). Shaded regions denote the standard error of mean across dRs and recordings. Twenty-eight dR configurations (3 × 2 up to 16 × 16, corresponding to 6-256 total discretized regions and areas of 1.94-0.046 mm$^2$) were investigated. **B)** Same as panel (A) for the pD residual impulse response (Res-IR) measure. **C)** Same as panel (A) for the pD residual periodicity (Res-Prd) measure. **D)** Same as panel (A) for the impulse response function settling rate (IRF-SR) measure. **E)** Same as panel (A) for the autocorrelation function impulse response (ACF-IR) measure. **F)** Same as panel (A) for the autocorrelation function periodicity (ACF-Prd) measure. **G)** Optimum dR yielding the largest mean stimulation-induced change (ΔIaM) for RNS Paradigm 1. **H)** Same as panel (G) for RNS Paradigm 2. **I)** Same as panel (G) for RNS Paradigm 3.

---

**Table S1.** Patients' clinical biographies and assessments.

A Optimum total discretized regions (TdRs)

| | LC | ACF-IR | ACF-Prd | IRF-SR | Res-IR | Res-Prd |
|---|---|---|---|---|---|---|
| Paradigm 1 | 25 | 20 | 9 | 20 | 36 | 6 |
| Paradigm 2 | 36 | 20 | 9 | 36 | 42 | 20 |
| Paradigm 3 | 20 | 9 | 20 | 30 | 49 | 49 |

B AUC-Values (Comparing RNS-ON and RNS-OFF means at optimum TdRs)

| | LC | ACF-IR | ACF-Prd | IRF-SR | Res-IR | Res-Prd |
|---|---|---|---|---|---|---|
| Paradigm 1 | 0.75 | 0.88 | 0.97 | 1.00 | 1.00 | 0.94 |
| Paradigm 2 | 0.63 | 1.00 | 0.78 | 0.69 | 0.94 | 0.75 |
| Paradigm 3 | 0.50 | 1.00 | 0.93 | 1.00 | 0.94 | 0.81 |

C P-Values (Comparing RNS-ON and RNS-OFF means at optimum TdRs)

| | LC | ACF-IR | ACF-Prd | IRF-SR | Res-IR | Res-Prd |
|---|---|---|---|---|---|---|
| Paradigm 1 | 1.62e-2 | 3.66e-07 | 5.58e-06 | 5.04e-07 | 4.72e-04 | 3.10e-02 |
| Paradigm 2 | 6.61e-04 | 6.00e-15 | 6.91e-04 | 4.02e-03 | 7.33e-05 | 1.29e-02 |
| Paradigm 3 | 5.53e-04 | 5.88e-10 | 5.67e-06 | 1.92e-04 | 1.84e-02 | 2.06e-02 |

---

**Table S2. RNS paradigms optimum total discretized regions (TdRs), AUC values and statistics for activity markers. A)** Optimum total discretized regions (TdRs) for each interictal activity marker (IaM) across RNS Paradigm 1-3. **B)** The areas under the ROC curves (AUC) for each IaM across RNS Paradigm 1-3. **C)** P-value statistics for each IaM when comparing means (averaged across recordings and dRs at the optimum TdR) of the IaMs during RNS pulses-ON and pulses-OFF periods.

---

**Movie S1. Dynamic statistical parametric mapping (dSPM) of RNS-evoked hemodynamic responses.** Movie shows localized cortical regions with significantly increased (reddish) and decreased (blueish) in CBV at each sliding window step, relative to reference activity (last 5 s of pulses-OFF periods; two-sided *Student's t-test*, $p < 0.01$, FDR corrected across pixels) for P8, RNS Paradigm 3. The dSPM is derived using a sliding window (5 s data-frames) at 1 s steps, by performing a pixelwise comparison of the mean activity at each window-step to the reference activity, beginning with the reference activity period, and sliding across the pulses-ON and pulses-OFF periods. The dSPM is overlaid on a mean baseline pD grayscale image.
